# Pan-cancer tumour classification and risk stratification from whole-genome somatic variants via dual-task representation learning

**DOI:** 10.64898/2026.03.02.26347318

**Authors:** Prima Sanjaya, Esa Pitkänen

## Abstract

Tumour typing from whole-genome sequencing is increasingly accurate, yet molecular subtyping from somatic variants remains challenging because of tumour heterogeneity and inconsistent clinical annotations. Here, we present Mutation-Attention Dual-Task (MuAt2), a Transformer model that jointly classifies histological tumour types and subtypes directly from somatic single-nucleotide variants, indels and structural variants. MuAt2 leverages encoders pre-trained on 2,587 pan-cancer whole genomes, and subsequently fine-tuned and evaluated on 14,527 tumour whole genomes from Genomics England spanning 15 tumour types and 68 subtypes. MuAt2 outperformed aggregated-feature deep baselines and conventional machine learning models. Fine-tuning improved both accuracy and calibration across independent cohorts processed with heterogeneous variant-calling pipelines. MuAt2 embeddings organised tumours by lineage and oncogenic processes, captured molecular subtype-defining driver events and improved prognostic stratification in gliomas. Finally, MuAt2 facilitated interpretation of metastatic tumours and cancers of unknown primary by inferring plausible tissue origins from somatic variant patterns. In conclusion, MuAt2 provides a transferable and interpretable modelling framework for cancer diagnosis and prognosis directly from whole-genome somatic variation.

## Introduction

Precision oncology seeks to stratify tumours based on their molecular characteristics in order to guide therapeutic decision-making [1]. Targeted interventions [2–4] have substantially improved response rates and survival compared with historical treatment regimens [5–8]. However, durable responses remain limited by intra- and intertumoral heterogeneity driven by clonal evolution and multi-scale genomic alterations ranging from single-nucleotide variants (SNVs) to large structural rearrangements (SVs) [9, 10]. This challenge is particularly pronounced in cancers of unknown primary (CUP), where the inability to resolve tumour lineage constrains evidence-based therapy selection.

The adoption of next-generation sequencing in research and clinics has created an opportunity to infer tumour identity directly from somatic variant patterns. Somatic variants reflect the evolutionary history of a tumour as well as tissue- and exposure-specific mutational processes [11–13], providing a rich substrate for computational modelling of tumour types and subtypes. Translating these signals into robust and scalable classification systems, however, remains a significant challenge, particularly when models must operate under realistic clinical infrastructure constraints such as limited compute capability.

Existing approaches for tumour classification based on somatic variants broadly fall into two categories: methods that aggregate variant-level features into summary statistics and models that explicitly represent individual variants. Aggregation-based approaches, including deep neural networks trained on mutation spectra derived from WGS data [14], demonstrated the feasibility of tumour typing from somatic variant profiles. More recently, attention-based architectures have enabled direct modelling of individual variants, improving performance across WGS and whole-exome sequencing (WES) datasets [15, 16]. Notably, passenger mutation patterns frequently encode stronger tissue-of-origin signal than sparse driver mutations, although driver-focused panel sequencing approaches have also achieved high classification accuracy [17]. Complementary approaches have leveraged chromosomal instability and copy number features in tumour typing [18].

Despite these advances, several limitations remain. Most existing approaches focus on either tumour type classification or unsupervised subtype discovery [19–21], rather than jointly modelling both tasks within a unified framework. Compared to tumour typing, identifying molecular subtypes remains more challenging, largely because subtype annotations are limited, inconsistent, or cohort-specific, constraining supervised learning approaches. In such settings, unsupervised methods are commonly employed to identify latent molecular structure that is subsequently interpreted as candidate subtypes [19–21]. Notably, our prior work demonstrated that a model trained under tumour type supervision (MuAt) nonetheless learned representations that encode subtype-discriminative information, suggesting that subtype-relevant signal is inherently embedded within somatic variant patterns [15]. A unified framework capable of robustly modelling tumour type and subtype from somatic variant data, while remaining computationally efficient and deployable in controlled, resource-limited environments, remains lacking.

In this study, we extend MuAt into a unified multi-task framework (MuAt2) engineered to jointly model tumour type and subtype directly from whole-genome somatic variant data. We train and evaluate MuAt2 on a large-scale cohort of 14,527 cancers from Genomics England (GEL) [22]. The resulting framework enables supervised learning of tumour type and subtype within a unified architecture, while remaining computationally efficient and compatible with Secure Processing Environments (SPEs). Through systematic benchmarking, we demonstrate that MuAt2 achieves robust classification performance and that its learned representations capture biologically meaningful structure, including associations with driver mutations and subtype-specific molecular variation, and prognostic information. Together, these results establish MuAt2 as a scalable genomic classification framework suitable for deployment in cancer clinics.

## Results

### Dual-task supervised learning of tumour types and subtypes with MuAt2

We developed a deep neural network model, Mutation-Attention Dual-Task (MuAt2), building on our earlier model, MuAt [15] (Fig. 1). MuAt2 is a Transformer model [23] that considers individual somatic variants observed in a tumour, and represents them by their variant type, position and other attributes. The primary distinction between the previous MuAt model and MuAt2 is the ability of the latter to classify both tumour types and subtypes at the same time given a set of somatic variants as the input. This is achieved by incorporating a separate classification path for both types and subtypes after a Transformer encoder (Methods). Joint optimisation over tumour type and subtype encourages a shared tumour representation that exploits the hierarchical relationship between labels and acts as an inductive bias that can improve generalisation.

**Figure 1:**
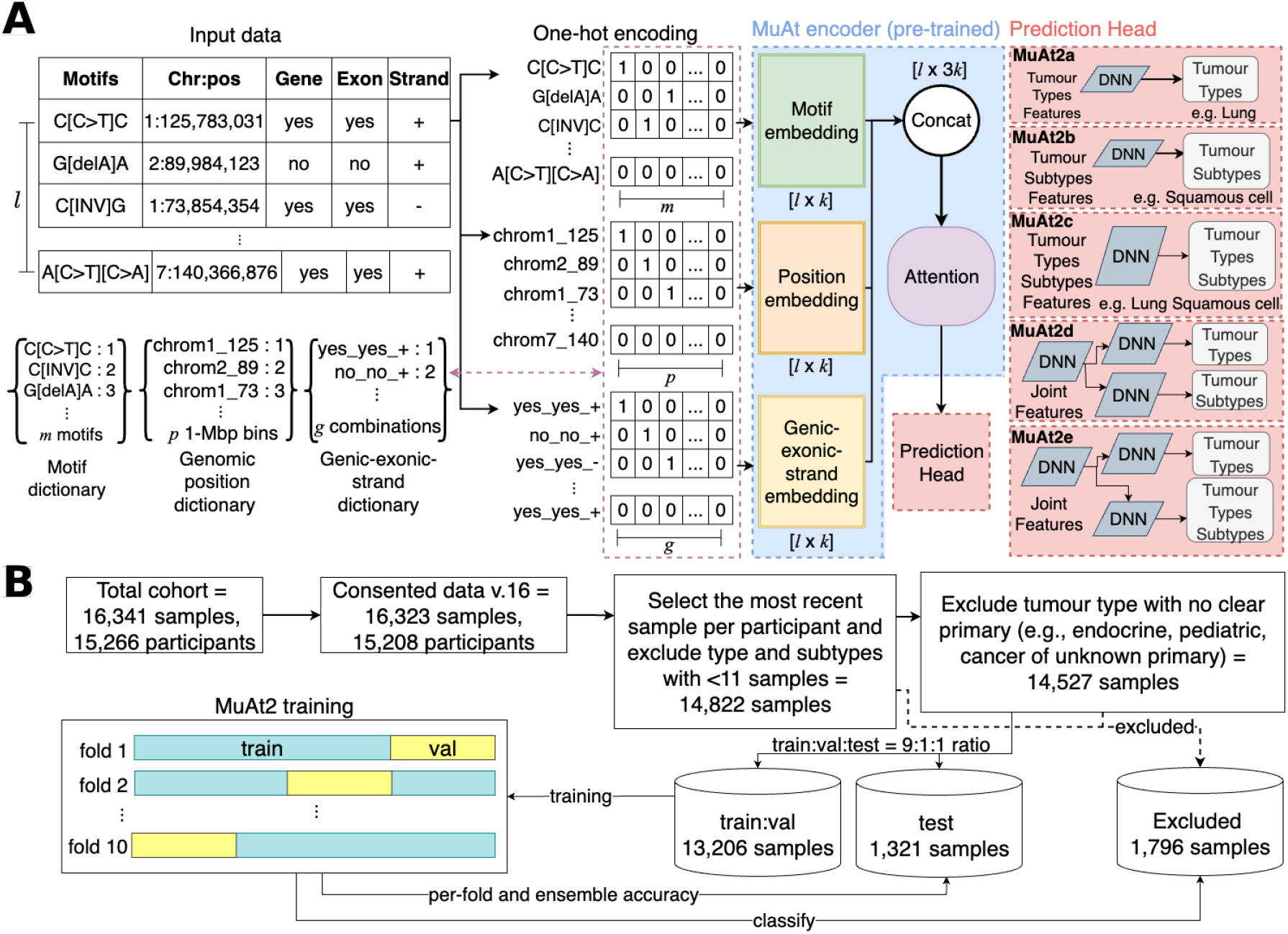
Overview of MuAt2 architecture and training procedure. (**a**) Schematic of the MuAt2 deep neural network model. Input somatic variants are encoded as by their sequence context (motif), genomic position, and genic–exonic–strand features, which are one-hot encoded, embedded, and concatenated. An attention module integrates these multi-view embeddings, and the resulting latent representation is passed to a prediction head. Different classification heads were tested: single-task models for tumour types (*e*.*g*., “lung”; **MuAt2a**), tumour subtypes (*e*.*g*., “squamous cell”; **MuAt2b**), or combined type–subtype labels (**MuAt2c**) and dual-task models jointly predicting tumour type and subtype (**MuAt2d**) or tumour type and specific type–subtype labels (**MuAt2e**). (**b**) Data selection and training procedure. MuAt2 models were trained on 13,206 tumours and performance was evaluated on the test set of 1,321 tumours. Tumours excluded from MuAt2 model training (n=1,796) were used to further evaluate model performance.

For each somatic variant, MuAt2 encodes its three-base sequence context (*e*.*g*., Ap[C>T]pG, Tp[delC]pC), genomic position in 1-Mbp bins, and genome region annotations indicating whether the variant occurs in a gene or exon, and the coding strand orientation for genic variants. MuAt2 is able to support any variant defined at a base-pair resolution; in this study, we experiment with models trained with somatic single nucleotide variants (SNVs), insertions and deletions (indels) and structural variants (SVs).

We developed alternative MuAt2 models to address different classification tasks. First, we created baseline models that classify tumour types (*e.g*., “Breast”), subtypes (*e*.*g*., “Squamous cell”), and combined type-subtype labels (*e*.*g*., “Breast squamous cell”) (Fig. 1**a**, MuAt2 models **a, b, c**). Then, we compared these baseline models with models that utilize joint classification heads, which simultaneously classify tumour types and subtypes in a single forward pass (Fig. 1**a**, MuAt2 models **d, e**). We trained these models, and other machine learning (ML) models including random forest (RF), extreme gradient boosting (XGB) and a previously proposed deep neural network model (DNN; [14] on 14,527 whole tumour genomes from 22 tumour types and 145 histological subtypes (Fig. 1, Supplementary Fig. 1; Methods) available in Genomics England [22, 24].

### Tumour type and subtype classification performance

We first evaluated the effect of classification head choice to tumour type and subtype performance when classifying a set of tumours not used in training. In all tasks, MuAt2 models with a joint classification head (MuAt2d and MuAt2e) achieved higher performance compared to the single-head baseline models (MuAt2a, MuAt2b, MuAt2c) (Fig. 2a, Supplementary Table 1). The best performance was achieved when utilizing somatic SNVs, indels and SVs annotated with genomic positions, resulting in accuracies ranging from 85.5% to 86.6% across individual cross-validation folds (per-fold accuracy) and 88.8% when combining predictions from all folds (ensemble accuracy) for tumour typing, 56.1%–59.1% per-fold (61.9% ensemble) for subtyping, and 55.4%–57.9% per-fold (59.1% ensemble) for type-subtype labeling. Similarly to our earlier work [15], performance was markedly improved when annotating variants with genomic positions, with additional covariates yielding smaller improvements.

**Figure 2:**
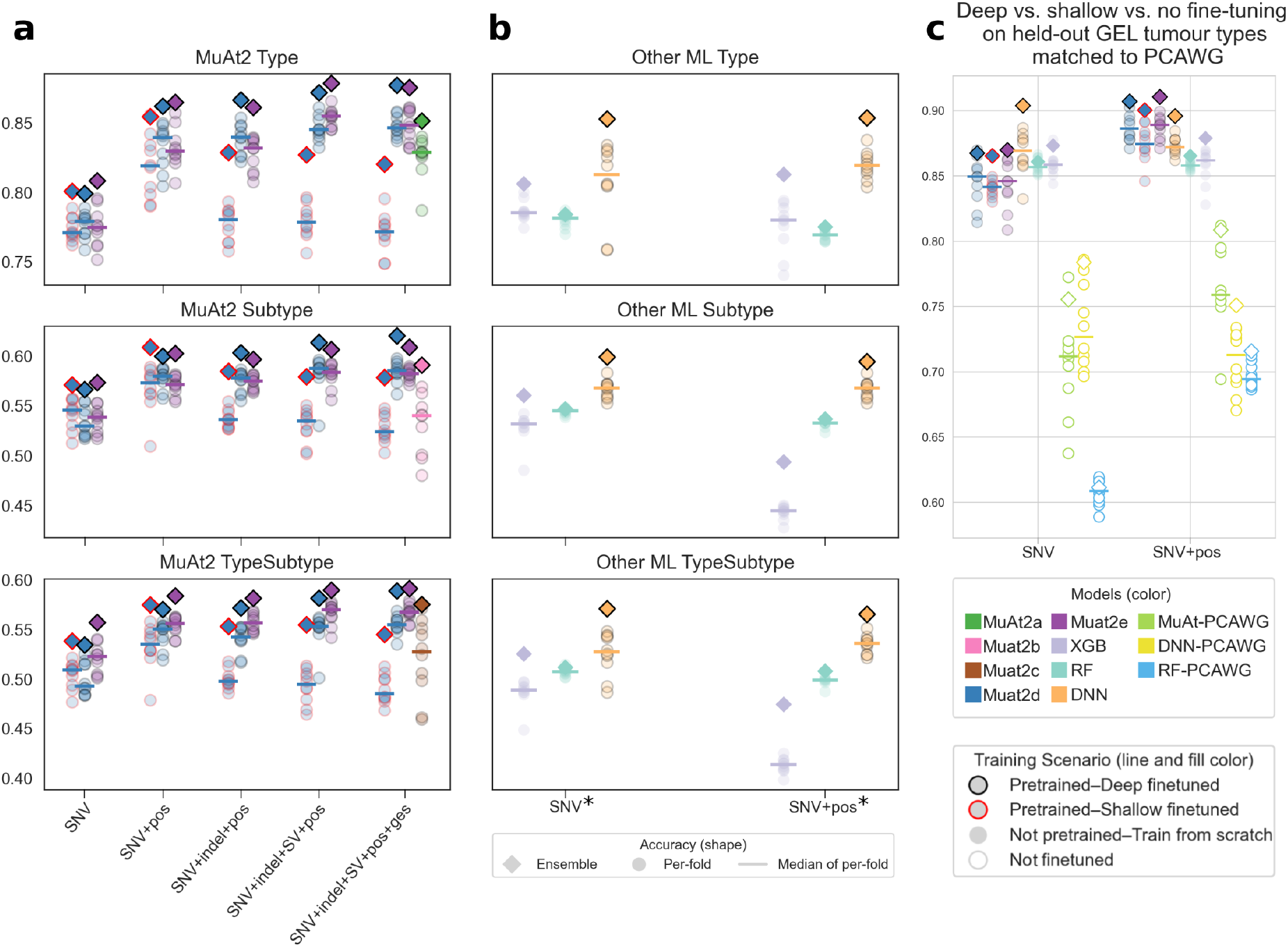
Performance of MuAt2 and other machine learning models in classifying tumour type, subtype and type-subtype labels in the Genomics England dataset. **a**) Accuracies of MuAt2 models trained with different variant types and annotations. Cross-validation per-fold, median of per-fold and ensemble accuracies are shown as points, lines and diamonds, respectively. **b**) Accuracies of DNN [14], random forest (RF), and extreme gradient boosting model (XGB) models. SNV* indicates the aggregated feature set used in [14]. **c**) Classification accuracies of deep fine-tuned models against shallow and no fine-tuned models on the seven tumour types those are found both in GEL and PCAWG datasets.

We then benchmarked MuAt2 against other machine learning methods. Dual-task models (MuAt2d and MuAt2e) yielded better performance than the other methods on all tasks (Fig. 2b, Supplementary Table 1). DNN [14] using an extended set of SNV features (Methods) performed similarly to MuAt models integrating SNVs and genomic positions (accuracy 76%–83%). Both RF and XGB failed to reach MuAt2 level of performance.

### Effective model portability across cohorts requires fine-tuning

We then sought to understand whether previously trained MuAt models can be reused with new data to facilitate deploying the models to clinical use. To this end, we used MuAt models pretrained on Pan-Cancer Analysis of Whole Genomes (PCAWG; [15]) data as MuAt2 encoders, and evaluated their performance on the Genomics England dataset. Given that the PCAWG dataset is smaller than GEL, and differs in how somatic variants were identified, we hypothesized that distribution shifts especially in indel and SV frequencies between the two datasets could pose a challenge to a pre-trained encoder. We thus explored the effect of fine-tuning to improve performance. Two transfer learning strategies were evaluated: (1) shallow fine-tuning, where only the classification head is updated while the encoder pre-trained with PCAWG data remains frozen (MuAt2d), and (2) deep fine-tuning, where all parameters, including embeddings and attention layers, are updated on GEL data (MuAt2a–e). Performance was benchmarked against models that reused MuAt’s pre-trained encoder without any additional fine-tuning.

To enable direct comparison between the datasets, and to isolate the impact of fine-tuning, we focused on the seven tumour types present both in PCAWG and GEL: breast, colorectal, lung, prostate, renal and upper gastrointestinal cancers, and melanomas. When evaluated on held-out GEL test data, all models benefited from fine-tuning (Fig. 2c, Supplementary Table 1). Deep fine-tuning consistently outperformed pretrained models without fine-tuning, increasing ensemble accuracies from 81% (MuAt) to 92% for MuAt2, 75% to 90% for DNN, and 72% to 87% for RF. Shallow fine-tuning of MuAt2 reached ensemble accuracies of 87—91%, comparable to deep fine-tuning. Furthermore, fine-tuned models demonstrated better calibration than pretrained models without fine-tuning (Supplementary Fig 2).

To assess whether MuAt2 retained generalisability after fine-tuning, we also tested it on a PCAWG dataset which had not been used in MuAt2 training including pre-training steps. The model achieved high performance (Supplementary Fig. 3) compatible to our previous results [15]. For instance, MuAt2 classified glioblastomas as adult gliomas with 90% accuracy, and haematological cancers with >90% accuracy. Upper gastrointestinal tumours were classified either as oesophageal or stomach adenocarcinomas, reflecting the tumour type labelling differences between the two datasets.

### Metastasis-driven misclassification in tumour type prediction

We next evaluated MuAt2’s classification performance across individual tumour types, using the MuAt2e dual-task ensemble model trained on somatic SNVs, indels and structural variants annotated with genomic positions and features. The model demonstrated strong and consistent performance across multiple cancer types (Fig. 3). Notably, classification of colorectal and breast cancers was highly accurate (97% and 95% accuracy, respectively). High performance was reflected in a two-dimensional UMAP projection of MuAt2 features, showing tumour type specific clustering (Fig. 4).

**Figure 3:**
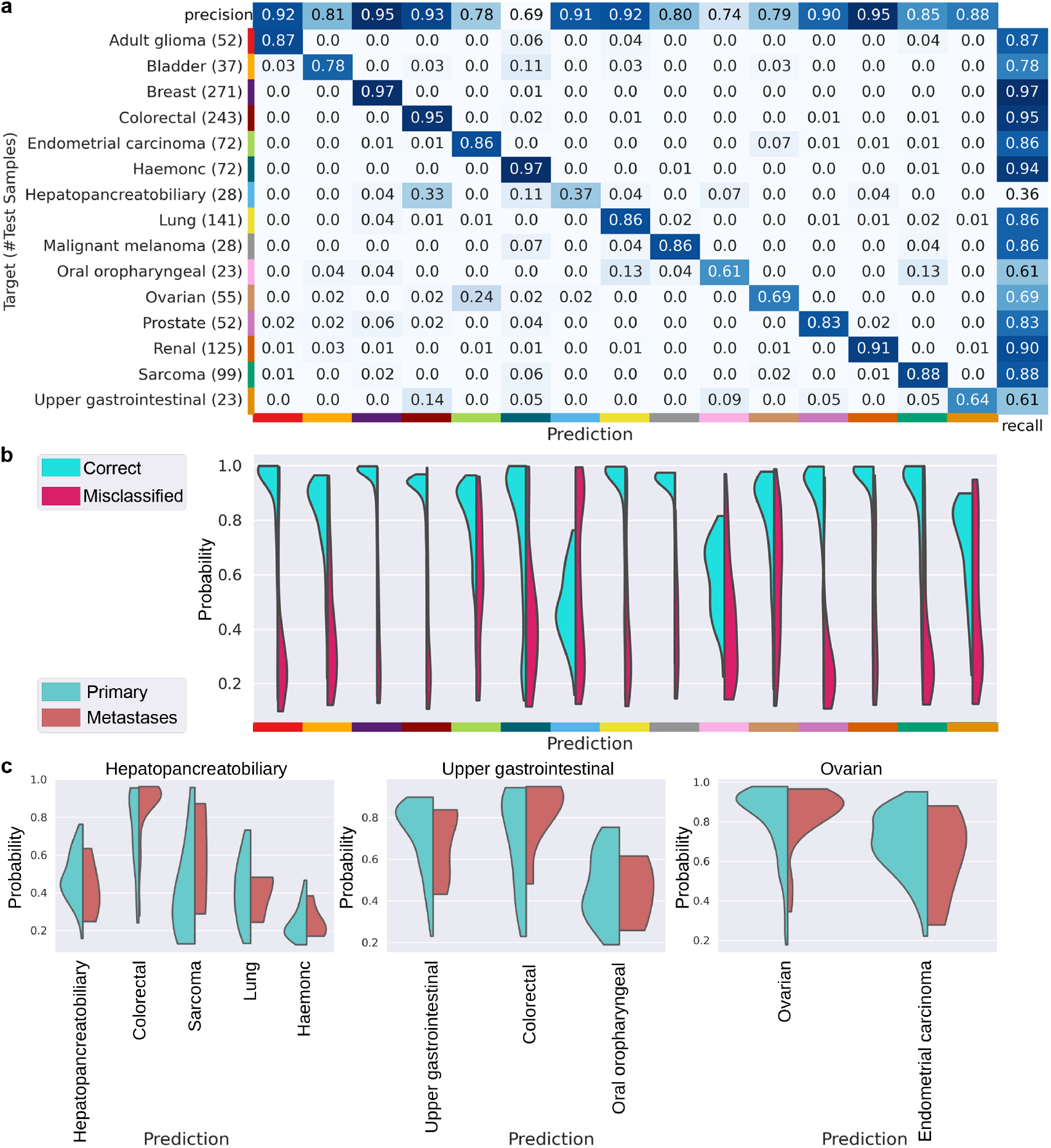
Evaluation of cancer type and subtype classification performance of MuAt2. **a**) Confusion matrix illustrating precision and recall for each cancer type on the test set. **b**) Top-1 probability distributions with the correct (blue) and misclassified cases (red) across cancer types reflecting the model’s confidence in tumour classification. **c**) Classification probabilities for cancers labelled as hepatopancreatobiliary, upper gastrointestinal or ovarian cancers stratified into primary (blue) and metastatic cancers (red).

**Figure 4:**
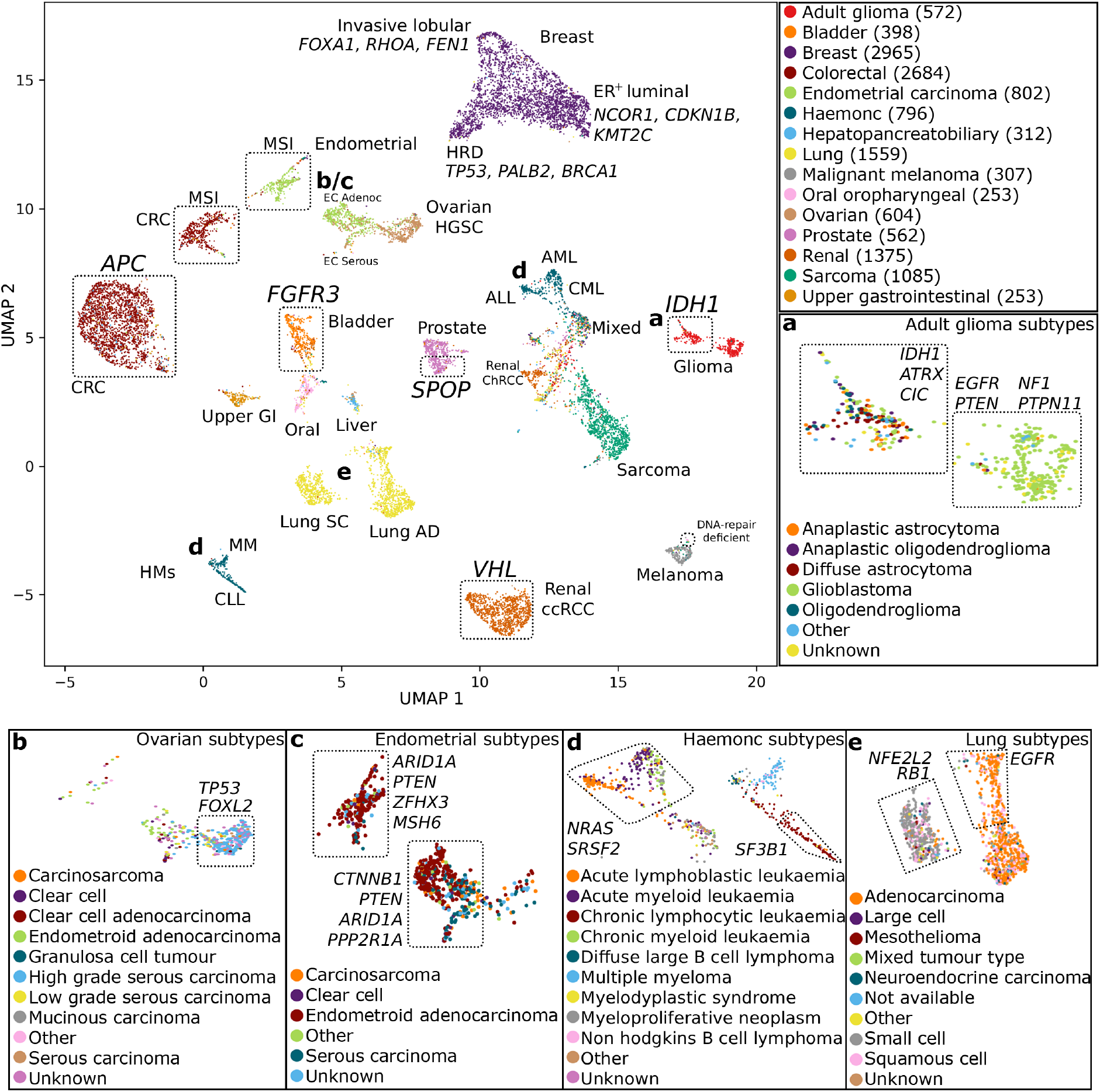
UMAP projection of MuAt2 tumour-level features in 14,527 GEL tumors. Annotations indicate tumor types and subtypes (*e*.*g*., lung adenocarcinoma, AD) and somatic alterations in cancer driver genes (*e*.*g*., *SPOP, APC*). Bold letters **a**–**e** correspond to panels showing tumours of a specific tumour type by subtype. The legend shows the number of tumours per each type. **a**) Adult gliomas clustered into glioblastoma and non-glioblastoma groups. **b**) In ovarian cancers, high grade serous carcinomas (HGSC) were clustered separately from the other tumours. The HGSC cluster was enriched for *TP53* and *FOXL2* alterations, the latter being a known driver in granulosa cell tumours. **c**) Endometrial adenocarcinomas clustered into MSI (left) and microsatellite-stable (MSS; right) tumors. Both **d**) haematological malignancies and **e**) lung cancers clustered into two groups with characteristic drivers. Note that panel **d** shows together two distinct clusters of HMs located apart on the main UMAP.

Performance declined for tumour types with non-specific mutational profiles, however. For instance, 24% of ovarian cancers were misclassified as endometrial, 13% of oral or oropharyngeal cancers as lung, and 14% of upper gastrointestinal tumours as colorectal. Across all tumour types, those labelled as hepatopancreatobiliary cancers were the most challenging to classify (37% accuracy). To investigate the origin of these errors, we examined the distribution of classification probabilities (Fig. 3b). Notably, several tumour types, particularly hepatopancreatobiliary and upper gastrointestinal cancers, were frequently assigned high classification probabilities despite being incorrectly predicted.

In order to better understand this behaviour, we examined top-1 classification probabilities by tumour type, stratified by whether the samples were labelled as primary or metastatic (Fig. 3c; Supplementary Fig. 4). We observed that tumours labelled as metastatic were sometimes misclassified with high confidence. For instance, many hepatopancreatobiliary and upper gastrointestinal tumours labelled as metastatic were confidently predicted as colorectal (Fig. 3c; Supplementary Fig. 5), compatible with the liver’s role as a common metastatic destination [25]. However, misclassified metastatic ovarian tumours also received similarly high probabilities compared to primary tumours likely due to the substantial overlap in the mutational profiles between ovarian and endometrial cancers. When we excluded metastatic tumours from the test set, classification accuracy improved notably: hepatopancreatobiliary accuracy increased from 37% to 50%, ovarian from 57% to 66%, and upper gastrointestinal from 58% to 74% (Supplementary Fig. 6).

### Lineage resolution within the haematopoietic hierarchy

While MuAt2 achieved strong performance in tumour type classification overall, haematological malignancies (HMs) present unique challenges due to their developmental diversity and overlapping genomic features. Within the GEL dataset, haematological malignancies are well represented and span multiple malignancy types, such as acute myeloid leukaemia (AML), acute lymphoblastic leukaemia (ALL), chronic myeloid leukaemia (CML), and mature B-cell neoplasms, coded as tumour subtypes in GEL. MuAt2 performed well on HMs with clear molecular and histological signatures. For instance, for ALL and AML, it achieved accuracy of 86% and 73%, respectively (Supplementary Fig. 8). However, performance was strongly influenced by class imbalance: the model tended to favour HMs with larger sample sizes, with reduced sensitivity in underrepresented HMs (Supplementary Fig. 7).

Misclassification patterns were particularly notable among haematological lineages. Tumours derived from progenitor stem cells in the bone marrow, such as ALL, AML, and CML, were frequently misclassified as another malignancy type of the same lineage. Despite high performance in ALL (true positive rate, TPR 86%) and AML (73%), 9% of ALL cases were misclassified as AML (12% vice versa), and 53% of CML cases misclassified as AML. These errors likely arise from overlapping mutational profiles due to shared developmental origins in early hematopoiesis. A similar pattern emerged among subtypes derived from more mature lymphoid or plasma cells. Non-Hodgkin B-cell lymphoma (NHBCL), diffuse large B-cell lymphoma (DLBCL) and multiple myeloma (MM) were frequently misclassified as CLL, with CLL receiving over 20% false positive predictions from these subtypes (Supplementary Fig. 8). These findings highlight the need for more balanced training data to improve classification performance.

### MuAt2 representations capture oncogenic and lineage-specific mutational programs

We next asked whether MuAt2 could resolve molecular tumour subtypes beyond histological classification without explicit supervision. We therefore assessed enrichment of cancer driver events across MuAt2-defined tumour clusters (Fig. 4) and quantified the extent to which driver alterations explained tumour-level mutational patterns (Fig. 5; Methods). To minimise confounding by extreme mutational processes, we excluded tumours with high tumour mutation burden (TMB), microsatellite instability (MSI), or *POLE* ultramutability from enrichment analyses.

**Figure 5:**
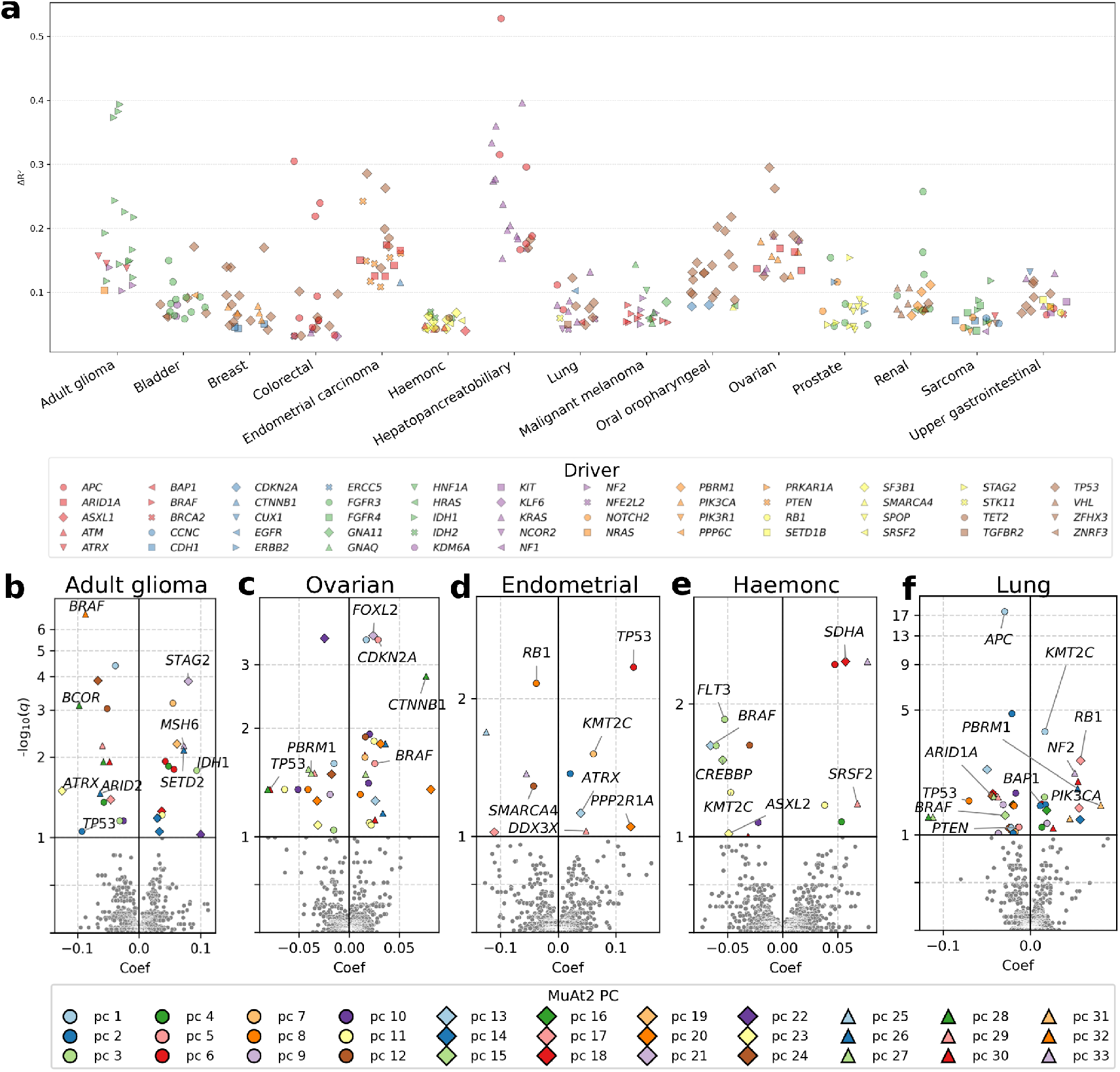
Association of MuAt2 feature principal components (PCs) and cancer driver events after excluding MSI, POLE, and high-TMB tumours. **a**) Performance gain (Δ*R*^2^) in predicting MuAt2 PCs when including the driver event (*i*.*e*., activating mutations in oncogenes or truncating, frameshift, or stop-gain mutations in tumour suppressor genes according to the Cancer Gene Census [26]) as a predictor in a least-squares linear model compared with a baseline model including only patient age and sex. Note that the same driver may appear more than once if it associates with multiple MuAt2 PCs. **b-f**) Volcano plots of coefficients *β* (x-axis) and − log_10_(*q*) values (y-axis) for association of MuAt2 PCs and driver events for **b**) adult gliomas, and **c**) ovarian, **d**) endometrial, **e**) haematological, and **f**) lung cancers. Only the association with highest − log_10_(*q*) is labelled.

Across cancer types, MuAt2 clusters were strongly enriched for known driver alterations, indicating that tumour-level mutational patterns capture underlying oncogenic and DNA repair processes (Fig. 4a–e, Fig. 5a–f, Supplementary Tables 2, 3). In particular, MuAt2 consistently separated tumours according to defects in DNA repair and replication fidelity. MSI cancers formed distinct clusters across colorectal and endometrial tumours (Fig. 4c), while homologous recombination repair–deficient breast cancers with *TP53, PALB2* and *BRCA1* mutations clustered together. When hypermutable tumours were included in the analysis, additional associations emerged with genes involved in DNA replication and repair, such as *BLM*, and with recurrent MSI target genes including *ACVR2A* and *TGFBR2* (Supplementary Fig. 14).

Beyond canonical repair pathways, MuAt2 features were associated with driver events linked to replication stress, chromatin regulation, and genome maintenance across tumour types (FDR<1%; Supplementary Fig. 15). These included established DNA repair and chromatin regulators (*TP53, ATRX, ARID1A*), as well as oncogenes and tumour suppressors with emerging or contextdependent roles in mutagenesis (*KRAS, NF1, PTEN, PIK3CA*). Such associations indicate that MuAt2 captures mutational consequences of diverse oncogenic states.

MuAt2 also resolved molecular subtypes defined by lineage- and differentiation-specific drivers. For instance, in adult gliomas, MuAt2 separated *IDH* -wildtype glioblastomas enriched for *EGFR* and *PTEN* alterations from *IDH1* -mutant gliomas frequently harbouring *ATRX* mutations (Fig. 4a). Similarly, renal, bladder, and prostate tumours clustered according to subtype-defining drivers such as *VHL, FGFR3*, and *SPOP*, reflecting known molecular stratifications.

Distinct mutational patterns were associated with developmental origin and cellular lineage. Haematological malignancies separated into clusters corresponding to progenitor-derived leukaemias versus mature B-cell and plasma-cell neoplasms (Fig. 4d), with associations to recurrent drivers including *FLT3, SRSF2, KMT2C* and *CREBBP*. In lung cancer, MuAt2 distinguished squamous and small-cell tumours enriched for *NFE2L2* /*KEAP1* activation and *RB1* loss from adenocarcinomas, within which *EGFR*-mutant tumours formed a distinct group (Fig. 4e). Rare *BAP1* alterations were associated with elevated mutation burden and distinct mutational patterns, consistent with impaired DNA repair.

Together, these results demonstrate that MuAt2 captures biologically meaningful mutational processes driven by DNA repair deficiency, oncogenic stress, and lineage identity, enabling unsupervised resolution of molecular tumour subtypes across diverse cancer types.

### MuAt2 captures independent prognostic information in adult gliomas

To assess whether MuAt2 representations capture prognostic information, we evaluated the association between MuAt2 feature PCs and overall survival in adult glioma patients using Cox proportional hazards models. Risk scores were derived from models trained using clinical covariates (*e*.*g*., age, sex, and grade), mutational signatures, MuAt2 PCs, and their combinations, and patients were stratified into equal-sized risk groups (Methods).

MuAt2 features improved prognostic discrimination beyond established clinical and genomic predictors. Adding MuAt2 features to clinical covariates increased the concordance index (*C*) from 0.781 to 0.810 (Δ*C*=0.029, 95% bootstrap CI 0.015–0.044; bootstrap *p <* 0.001; likelihoodratio test (LRT) *p* = 5.67 *×* 10^−7^). Similarly, adding MuAt2 to mutational signatures improved performance (*C*=0.791 to 0.807; Δ*C*=0.016, 95% bootstrap CI 0.005–0.026; bootstrap *p* = 0.001; LRT *p* = 0.006). When combined with both clinical and mutational signature variables, MuAt2 further improved prognostic performance (*C*=0.818; Δ*C*=0.014, 95% bootstrap CI 0.004–0.024; bootstrap *p* = 0.005; LRT *p* = 0.02). Kaplan–Meier curves showed clear separation across equal-sized predicted risk groups for all models (Fig. 6a, Supplementary Fig. 23).

To examine the contribution of individual predictors, we inspected hazard ratios from multivariable Cox models. MuAt2 PCs retained independent prognostic associations after adjustment for clinical covariates and mutational signatures (Fig. 6b, Supplementary Table 12). Clinical covariates showed known associations with survival, including increased hazard with higher tumour grade and reduced hazard for *IDH1* -mutant and lower-grade gliomas. In combined models, MuAt2 components remained stable predictors.

**Figure 6:**
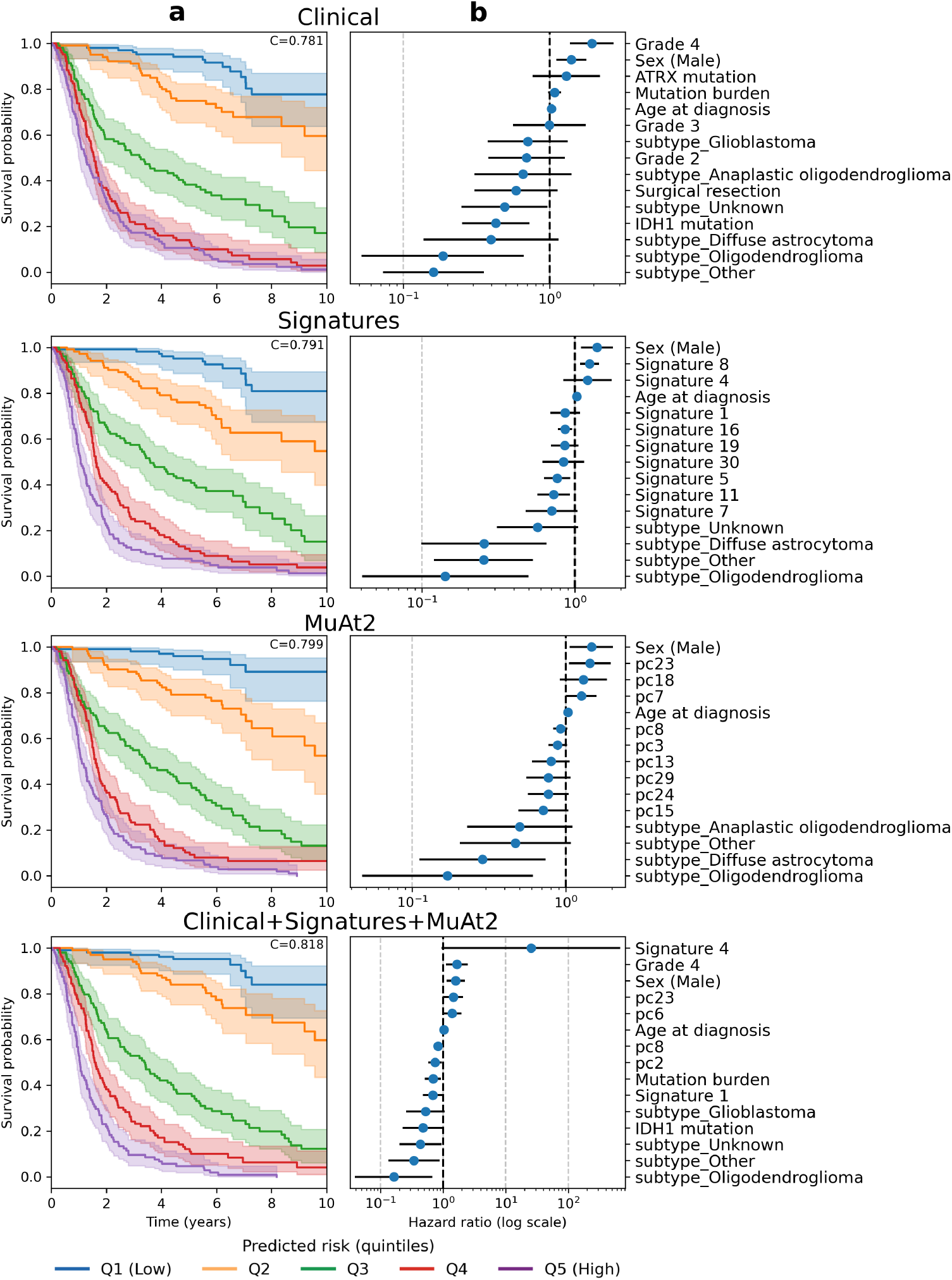
Prognostic stratification in adult gliomas (n=518) using four Cox models: clinical variables only, mutational signatures, MuAt2 features and a model combining these three sets of variables. Both mutational signature and MuAt2 models include age, sex, and the tumour subtype as variables. **a**) Kaplan–Meier curves showing survival stratified by predicted risk into equal-sized quintiles derived from the four Cox models. Performance quantified by the *C*-index is shown for each model. **b**) Forest plots showing multivariable hazard ratios for the top 15 variables ranked by *p*-value.

In glioblastoma, prognostic stratification was more challenging overall, yet MuAt2 improved risk discrimination over clinical covariates (*C* 0.705 vs 0.717; LRT *p*=0.008) but did not further improve performance over the combined clinical, driver, and signature model (*C* 0.704 vs 0.762; LRT *p*=0.191). Risk group separation was most apparent in models incorporating MuAt2 features (Supplementary Figs. 22, 24, and Supplementary Table 13).

### Prediction of likely tissue origin in metastatic cancers, and cancers with uncertain histology

To evaluate MuAt’s capability to provide clinical utility as a primary type predictor for tumours with non-specific histological annotation or unknown primary origin [27], we analysed a heldout subset of 1,796 tumour samples excluded from training (Methods). We stratified tumours into four groups: metastatic tumours with 1) specific and 2) non-specific histology (*e*.*g*., CUPs, and childhood and endocrine tumours), and primary tumours with 3) specific and 4) nonspecific histology. We then performed ensemble classifications with MuAt2e, and compared classifications with the tumour types assigned by the Genomic Laboratory Hubs (GLH) of the National Health Service, the tumour type groups curated by GEL, and the metastatic site corresponding to the biopsy or surgical sampling location (Methods).

Among metastatic tumours with clearly defined histology, MuAt2 predictions were largely consistent with both the GLH annotations and the GEL-curated tumour groups (Fig. 7a). Melanomas, and colorectal and lung adenocarcinomas showed near-complete agreement, and the predicted primary often aligned with the recorded metastatic site. We also observed several cases where GLH-assigned and GEL-curated types diverged, yet MuAt2 provided a high-confidence call that matched one of the labels. Discordances were mainly confined to low-confidence predictions, which typically reflected more ambiguous mutational profiles. In metastatic tumours with non-specific histology assigned by GLH, prediction confidence was generally lower, and high-confidence cases only partially aligned with the curated types (Fig. 7b).

**Figure 7:**
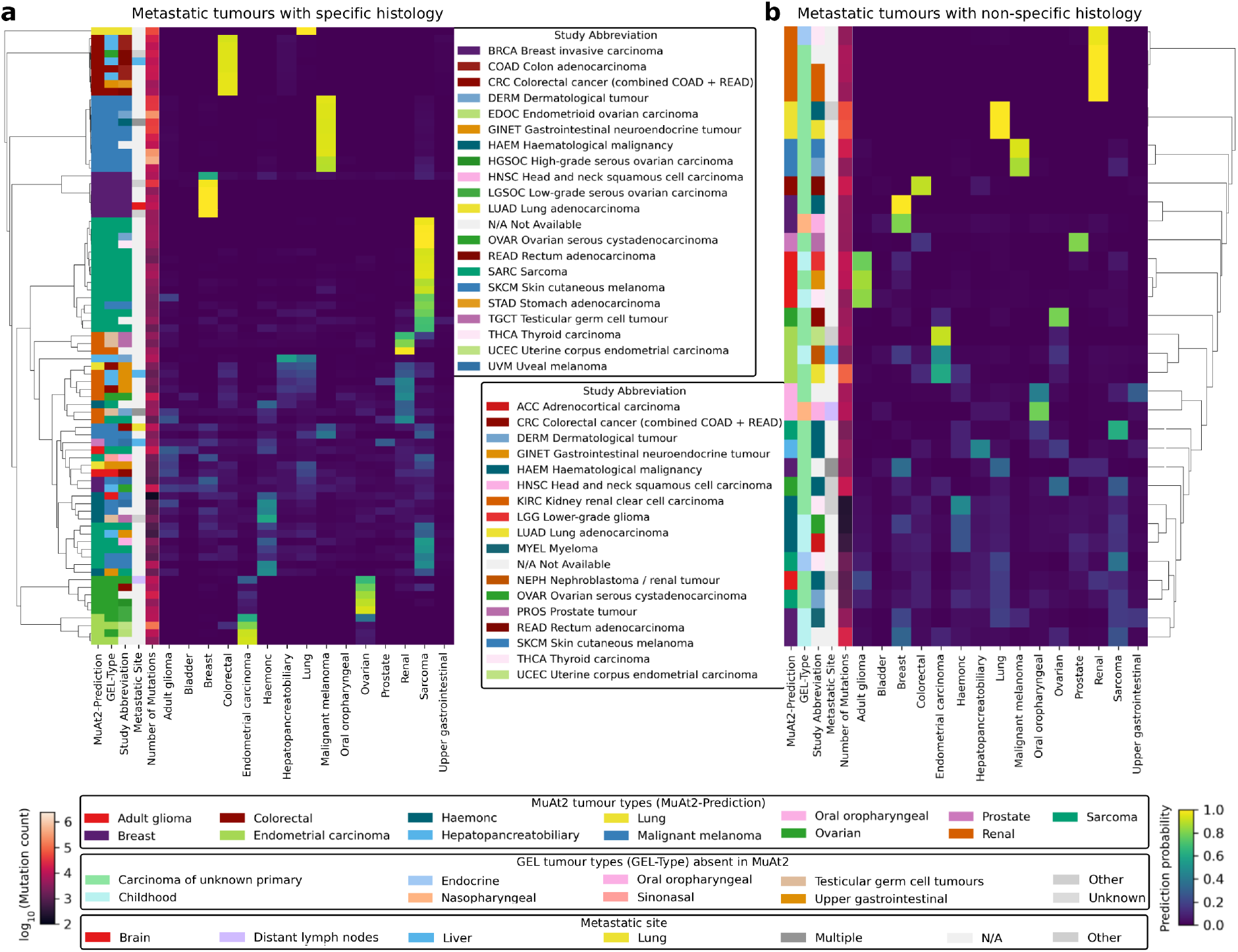
Ensemble predictions from the MuAt2e model for metastatic tumours stratified by histological clarity. Tumours were categorised by whether they had **a**) specific (*e*.*g*., breast, colorectal, bladder) or **b**) non-specific histology (*e*.*g*., CUP, childhood, nasopharyngeal, endocrine) based on GLH diagnosis. Heatmaps show MuAt2 prediction probabilities. Annotation columns show tumour type predicted by MuAt2, GLH diagnosis (“GEL-Type”), GEL-curated tumour type (“Study Abbreviation”), site of metastasis, and the number of mutations (log_10_-scale).

In primary tumours with non-specific histology, prediction confidence was low, and highconfidence calls showed only partial alignment with the curated labels (Supplementary Fig. 20). Notably, many paediatric brain tumours were classified as haematological cancers, likely due to their low mutation burden and the absence of paediatric brain tumours in our training dataset. Finally, in the group of primary tumours with well-defined histology, MuAt2 predictions were largely consistent with both the GLH annotations and the curated labels (Supplementary Fig. 21).

## Discussion

We introduced MuAt2, a dual-task machine learning framework to jointly predict histological tumour types and subtypes directly from whole-genome somatic variants. Trained on 14,527 tumours from Genomics England [22, 24], MuAt2 achieves strong tumour-type performance and substantially improved subtype resolution compared with single-task baselines and aggregated-feature classifiers [14, 15]. These results reinforce the notion that passenger-dominated mutation patterns encode robust tissue-of-origin signals [11, 12], and demonstrate that modelling individual variants with attention enables effective reuse of this information across diagnostic and prognostic tasks.

A central design feature of MuAt2 is the coupling of tumour type and subtype learning within a shared representation. Dual-task heads consistently outperformed single-task alternatives, indicating that jointly optimising related label hierarchies provides a useful inductive bias in cancer classification. Subtype prediction remains intrinsically more challenging than tumour typing, reflecting biological heterogeneity and annotation variability [19, 20], yet MuAt2 recovered subtype-relevant structure at pan-cancer scale. Importantly for clinical deployment, MuAt2 predictions were found to be well-calibrated.

We further show that model portability across cohorts requires fine-tuning. Direct reuse of PCAWG-pretrained encoders [15, 28] was suboptimal when applied to Genomics England data, consistent with distribution shifts arising from differences in cohort composition and variantcalling pipelines [29, 30]. Fine-tuning improved both accuracy and calibration, highlighting that effective transfer of genomic deep models depends on controlled adaptation rather than static reuse. This finding has direct implications for deployment in privacy-sensitive infrastructures such as national genomic initiatives [22, 24], and aligns with emerging paradigms in federated and distributed learning for medical AI [31].

Beyond classification performance, MuAt2 learns interpretable tumour embeddings structured by oncogenic processes and developmental origin. Embedding-based organisation of tumours aligned with canonical DNA repair phenotypes, including microsatellite instability and homologous recombination deficiency [12], and with lineage-defining driver alterations [26]. These results indicate that MuAt2 captures mutational consequences of genome instability, replication stress, and chromatin dysregulation rather than merely encoding tumour labels.

MuAt2 also provides informative signals in clinically challenging contexts. In adult gliomas, MuAt2-derived representations additionally captured prognostic information that was independent of established clinical variables and mutational signatures: adding MuAt2 features improved risk discrimination and retained significant associations in multivariable models.

MuAt2 demonstrated utility in clinically challenging settings. In adult gliomas, MuAt2-derived representations improved risk stratification over established clinical covariates and mutational signature exposures, indicating MuAt2 captures prognostic signals not fully reflected by conventional predictors. In metastatic tumours, high-confidence predictions frequently aligned with plausible primary origins and dissemination routes [32], consistent with the persistence of tissue-specific mutational patterns after metastatic spread. In contrast, lower-confidence outputs were enriched among tumours with non-specific histology or limited representation in training, highlighting the importance of comprehensive reference datasets. These observations suggest that mutation-level whole-genome models may complement histopathology in cancers of unknown primary and ambiguous metastatic presentations, while emphasising the need for prospective clinical validation.

Several limitations merit consideration. Subtype performance remains constrained by class imbalance and variability in clinical annotation, particularly for rare and paediatric entities. The current framework focuses on somatic SNVs, indels and structural variants, and does not integrate complementary subtype-defining modalities such as copy number alterations, methylation, transcriptomics or proteomics. Finally, although Genomics England provides a uniquely large and clinically curated cohort [22, 24], validation across additional national and international sequencing programmes will be necessary to establish robustness.

In summary, joint learning of tumour types and subtypes from somatic variant-level wholegenome representations improves performance relative to single-task and aggregated-feature approaches [14, 15], and yields embeddings structured by histological and molecular tumour identity. Our findings establish MuAt2 as a practical and scalable framework for tumour diagnostics and risk stratification, providing a foundation for developing genomic AI systems for clinical use.

## Materials and methods

### Evaluating transfer learning methods

We employ transfer learning to overcome computational limitations within the Secure Preprocessing Environment of Genomics England without significant GPU-resources, while leveraging pretrained models that were initially trained on a smaller dataset (PCAWG) [28].

MuAt2 encoder consists of three embedding layers followed by an attention layer, with all parameters initialised from MuAt pretrained on PCAWG data. Each embedding layer (motif, position, and annotation) transforms its respective input into embeddings representing the sequence context, genomic position, and annotations. To train MuAt2, we adopt two approaches: 1) updating only the classification head parameters while keeping the encoder frozen, and 2) performing deep fine-tuning by updating all parameters. We evaluate the performance of finetuned models with pretrained models without fine-tuning, evaluating on matching tumour types (PCAWG vs GEL) for direct comparison.

### Somatic variant data

We used the aggregated high-quality somatic variant callset (somAgg v0.2) of tumour whole genomes provided by the 100,000 Genomes Project, Genomics England [33]. This dataset is derived from 16,341 tumour whole-genome sequences from 15,226 individuals, spanning 22 cancer types and 145 histological subtypes (data release v.16) [22]. Each tumour sample in the dataset is paired with a corresponding matched normal tissue sample, and whole-genome sequenced to average depth of coverage of 100x and 30x, respectively. The dataset contains annotated SNVs and small indels (<50 bp) identified in a workflow including alignment using the iSAAC Aligner (version 03.16.02.19) and small variant calling with tumour-normal subtraction performed with Strelka2, with annotated somatic VCF files generated with CellBase [33]. All bioinformatics workflows utilized to create the somAgg dataset adhered to the Illumina North Star Version 4 Whole Genome Sequencing Workflow (NSV4, version 2.6.53.23) and were conducted with 150 bp paired-end reads on a single lane of an Illumina HiSeq X instrument, using the Homo sapiens NCBI GRCh38 assembly with decoys as the reference genome. We also used the set of somatic structural variants identified with MANTA.

### Data preprocessing

#### Somatic variant data preprocessing

We used only high-quality somatic variant calls that passed all quality checks (FILTER=PASS). Tumour type annotations were available in GEL metadata. Primary tumour types (GEL-Type) are based on clinical assignments provided by the GLH of the National Health Service and are subsequently reviewed and curated by GEL through an internal genomic review. In addition, GEL defines tumour type groupings (“Study Abbreviation”) that aggregate related primary tumour types for cohort organisation and downstream analyses. For samples labelled as metastatic, the anatomical site corresponding to the biopsy or surgical sampling location is recorded as the metastatic site and may differ from the assigned primary tumour type.

In the GEL v16 data release, informed patient consent was in place for 15,208 study cohort participants (16,323 out of 16,341 tumour samples, v16 release). As some participants had more than one tumour, we selected the most recent delivered tumour sample to represent each participant. The remaining samples from participants with multiple tumours were retained for joint analyses with non-specific histology and metastatic cases.

To enable stratified splits for training, validation and testing under 11-fold cross-validation, we excluded from training (i) tumour types with unclear histology (*e*.*g*., childhood cancers and cancers of unknown primary) and (ii) tumour type–subtype categories represented by fewer than 11 samples (*e*.*g*., breast adenocarcinoma and breast ductal carcinoma). In total, 14,527 samples across 15 tumour types and 68 subtypes (combined into 116 tumour type–subtype labels) were used for training, validation and testing, in 9:1:1 fractions, respectively (Fig. 1, Supplementary Fig. 1).

In total, we excluded 1,796 samples consisting of non-representative tumours from participants with multiple samples, undetermined types, and rare type–subtype labels annotated as primary or metastatic. To assess how well the model performs when histologic conclusions are uncertain, we split these samples on their metastatic status and whether the tumour types are well-defined. Out of these, 34 metastatic samples had unclear histology, 108 metastatic samples had clear histology, 315 primary samples had unclear histology, and 1,339 primary samples had clear histology. Complete information on the dataset is available in Supplementary Figs. 1, 18, and Supplementary Table 8.

#### Variant annotation and tokenisation

We followed the same preprocessing procedure as in our previous study [15], with sequence motifs, genomic positions, and variant annotations (*i*.*e*., genic-exonic-strand) each tokenised using the same predefined dictionaries in the encoder layer. For instance, suppose a single base substitution from C to T at position *p*, represented as [C *>* T]_*p*_. For instance, a C-to-T substitution within context ApCpG is written as *A*_*p*−1_[C *>* T]_*p*_*G*_*p*+1_ (*i*.*e*., A[C>T]G). The variant types include six substitution classes defined relative to the pyrimidine base, as well as insertions and deletions for each nucleotide (A, C, G, and T), structural variant breakpoints, and retrotransposon insertions. Genomic positions were encoded in 1 Mbp bins. For instance, the position chr1:2,213,687 falls within the range 2,000,000–2,999,999 and is encoded as 1_2, where ‘1’ indicates chromosome 1 and ‘2’ denotes the bin index for that 1 Mbp range. Somatic variants were further categorised based on their genomic context and strand relationship. Each variant was classified as occurring in a gene (“genic”) or exon (“exonic”) and assigned to one of the following strand categories: (1) same-strand with gene, (2) opposite-strand with gene, (3) overlapping two genes on opposite strands, or (4) intergenic. Lastly, as the GEL VCF files were provided in GRCh38/hg38, while the MuAt pretrained models used GRCh37/hg19 human genome reference, we converted the hg38 genomic position coordinates to hg19 using the Python package ‘liftover’. We used only autosomal variants, replicating the exact preprocessing steps used in MuAt. There were a total of 532,474,373 somatic variants, consisting of 348,008,311 single nucleotide variants (SNVs), 181,480,751 insertions and deletions (indels), and 2,985,311 structural variants (SVs), serves as training data for GEL (Supplementary Figure 18)

#### Data preprocessing for benchmarking ML models

To benchmark MuAt2 against other machine learning models, we preprocessed the inputs following the methodology described in [14] for Random Forest (RF), Deep Neural Network (DNN), and Extreme Gradient Boosting (XGB), all of which use aggregated data as input. Specifically, we aggregated 96 triplet motif counts per sample, consisting of SNVs with both flanking 5’ and 3’. These aggregated features were combined with mutational distribution features represent the counts within each 1-megabase bin spanning the genome. The total number of variant counts in each bin were normalised to the total number of the corresponding mutational events across the genome.

### Experimental Setup

#### Training, validation and testing of MuAt2 models

To train the MuAt2 models, we divided the data into 11 folds, stratified by tumour type and subtype, allocating 10 folds for cross-validation (9:1 training-validation ratio) and reserving the last fold for testing. Since the encoder hyperparameters had been optimised previously, we used the best set of hyperparameters which had been found during the optimisation of the pretrained models (see Supplementary Table 9). For the final classification, we ensembled the 10 models by averaging their probabilities and selecting the class with the highest score. The same splits were used for training, validation, and testing all models.

We trained MuAt2 to minimise the sum cross-entropy of tumour types (MuAt2a), tumour subtypes (MuAt2b), and tumour type subtypes (MuAt2c), as well as the combination of these (MuAt2d, MuAt2e). The loss functions for each of these are denoted as:

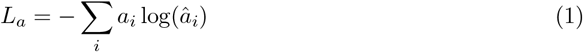

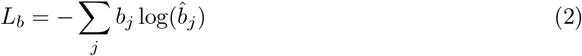

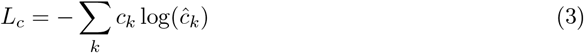

where *â*_*i*_, 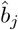, and *ĉ*_*k*_ are the predicted probabilities of tumour type *i*, subtype *j*, and type subtype *k. a*_*i*_, *b*_*j*_, *c*_*k*_ ∈ {0, 1} denote whether *i, j*, and *k* are the correct tumour type, subtype, and type subtype, respectively. MuAt2d loss is computed by summing type and subtype losses: *L*_*d*_ = *L*_*a*_ + *L*_*b*_, while MuAt2e is computed by summing type and type-subtype losses: *L*_*e*_ = *L*_*a*_ + *L*_*c*_. We experimented with no fine-tuning, shallow fine-tuning (by freezing the encoder and updating only the classification head parameters), and deep fine-tuning (*i*.*e*., updating all parameters), to assess the benefits of transfer learning and pretrained models.

#### Perfomance metrics

We report the performances using confusion matrix, with 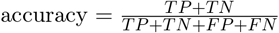, precision 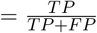, and 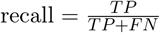, where TP, TN, FP, and FN denote the counts of true positives, true negatives, false positives, and false negatives, respectively.

### Association analysis of MuAt2 feature principal components

We combined the latent features from ten MuAt2e models inputing SNVs, indels and SVs annotated with positions and genomic features (*d* = 640 features) and reduced feature dimensionality using PCA retaining 95% of the variance (33 principal components). To interpret these PCs, we applied complementary association analyses addressing three questions: (i) whether individual driver events shift specific PCs, (ii) whether PCs capture broader mutational changes, and (iii) whether MuAt2-derived clusters are enriched for specific drivers.

Known cancer driver events were defined according to the Cancer Gene Census [26]. Samples were annotated as “driver-positive” when harbouring either hotspot mutations in oncogenes or truncating, frameshift or stop-gain mutations in tumour suppressor genes. Driver prevalences by tumour type are presented in Supplementary Fig. 19 and Supplementary Table 10.

For driver–PC associations, we fitted ordinary least-squares (OLS) regression models for each principal component within tumour-type–specific cohorts. A baseline model included only age and sex as covariates,

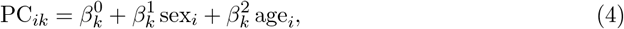

and a second model additionally included the driver event of interest,

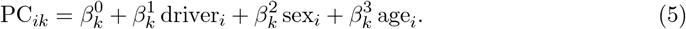

The change in explained variance (Δ*R*^2^) between the two models quantified the effect of the driver on each PC.

Associations between PCs and mutational burden or mutation-type counts were evaluated using analogous OLS regressions with log_10_(count + 1) as the outcome and PCs plus covariates as predictors:

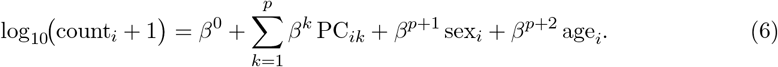

where *i* denotes the sample, *k* the PC, and *p* the number of PCs included as predictors (*p*=33).

We performed unsupervised clustering of MuAt2 principal components using *k*-means, with the number of clusters *C* selected by maximising the silhouette score. This first-level clustering produced *C* = 13 clusters (Supplementary Fig. 9). To explore potential substructure, *k*-means was re-applied within each primary cluster, again selecting *C* by the silhouette criterion, yielding 45 second-level clusters in total (Supplementary Fig. 10).

For each cluster at both levels, enrichment of driver events was assessed by comparing frequencies inside versus outside the cluster using one-sided Fisher’s exact tests (alternative = “greater”), reflecting a priori interest in enrichment rather than depletion (Supplementary Tables 2, 3).

As an auxiliary analysis, we modelled driver presence as the outcome in multivariable regressions fitted separately for each driver event *g*, using all MuAt2 PCs as predictors and adjusting for age, sex, and tumour type/subtype:

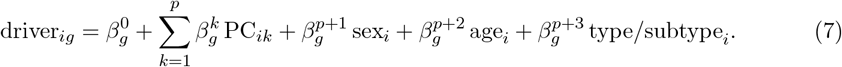

### Survival analysis

Associations with overall survival were evaluated using Cox proportional hazards models in adult glioma and glioblastoma cohorts. Models were fitted using clinical covariates (*i*.*e*., age, sex, mutation burden, subtype, surgical/biopsy, tumour grade, and mutation statuses of *IDH1, ATRX* and *TERT*), mutational signature exposures (COSMIC v2), MuAt2 principal components, and their combinations. In glioblastoma, driver variables were restricted to events observed in at least four samples to ensure stable estimation. Model discrimination was assessed using the concordance index (C-index). Improvements from adding MuAt2 were evaluated by bootstrap resampling of the C-index (1,000 resamples) and likelihood ratio tests comparing nested models. Δ*C* values represent mean bootstrap differences in concordance index between nested models with 95% confidence intervals obtained from the bootstrap distribution.

For visualisation, predicted risk scores were obtained from fitted models, and patients were ranked and stratified into equal-sized quintiles. Kaplan–Meier curves were generated for each risk group. Multivariable hazard ratios were extracted from combined Cox models and displayed for the top variables rank.

## Data Availability

All data from the National Genomic Research Library (NGRL) used in this research are available in the Genomics England Research Environment.

https://www.genomicsengland.co.uk/research

## Acknowledgements

We thank Dr. Mervi Aavikko, Dr. Outi Kilpivaara, Prof. Pirjo Laakkonen, Dr. Laura Langohr, and Prof. Ulla Wartiovaara-Kautto, Dr. Pia Vahteristo, and Siiri Reinikka for scientific discussions. We also thank Nora Schreiber and Daniyar Karabayev for technical assistance. We acknowledge funding from the Research Council of Finland (#322675), the Sigrid Jusélius Foundation, and the Cancer Foundation Finland. We gratefully acknowledge the participants of the National Genomic Research Library (NGRL), whose contributions made this research possible. Secure access to the NGRL under project ID 837 was provided by Genomics England, which delivers the NGRL in partnership with NHS England, and is wholly owned by the UK Department of Health and Social Care. The NGRL contains participants’ health data collected by the NHS as part of their care, along with samples and data from their participation in research, for which fully informed consent has been obtained. This includes genomic and clinical data provided through the NHS Genomic Medicine Service, as well as data obtained through research studies, including the 100,000 Genomes Project and the Generation Study, both of which are delivered in partnership with the NHS, and from other research cohorts involving external collaborators. The authors wish to acknowledge CSC – IT Center for Science, Finland, for additional computational resources.

## Data Access

Data from the National Genomic Research Library (NGRL) used in this research are available within the secure Genomics England Research Environment. Access to NGRL data is restricted to adhere to consent requirements and protect participant privacy. Data used in this research include: somatic aggregated variant call (somAgg), somatic SV, and clinical metadata as provided through the LabKey in Genomics England Research Environment. The overview of genomics data used in the research, release v.16 (13th October 2022) can be found at https://re-docs.genomicsengland.co.uk/release16/. Preprocessed data is available in the Research Environment at /re_gecip/machine_learning/muat/data.

Access to NGRL data is provided to approved researchers who are members of the Genomics England Research Network, subject to institutional access agreements and research project approval under participant-led governance. For more information on data access, visit: https://www.genomicsengland.co.uk/research

## Programming environment

MuAt2 was implemented in Python 3.7 using the PyTorch 1.8.0 deep learning framework. The Jiao *et al*. [14] model was evaluated using TensorFlow 2.0 in Python 3.6. Random forest models were implemented using scikit-learn 0.21.3, and gradient boosting models using xgboost 2.1.0. Statistical analyses were conducted using scipy 1.5.3 and statsmodels 0.12.1. Data processing and visualisation were performed using pandas 1.3.4, seaborn 0.11.2 and umap-learn 0.5.1. All models were trained on the HPC Double Helix CPU cluster (24 cores per node) within the Genomics England Research Environment.

## Code availability

Source code for MuAt2 is available at https://github.com/primasanjaya/mutation-attention. Due to data access restrictions of the Genomics England, trained models and parameter check-points are only available at /published_data_archive/RR837/muat2_checkpoint/ within the Genomics England Research Environment.

