## Supplementary figures for "Pan-cancer tumour classification and risk stratification from whole-genome somatic variants via dual-task representation learning"

[3]iCAN Digital Precision Cancer Medicine Flagship, Finland

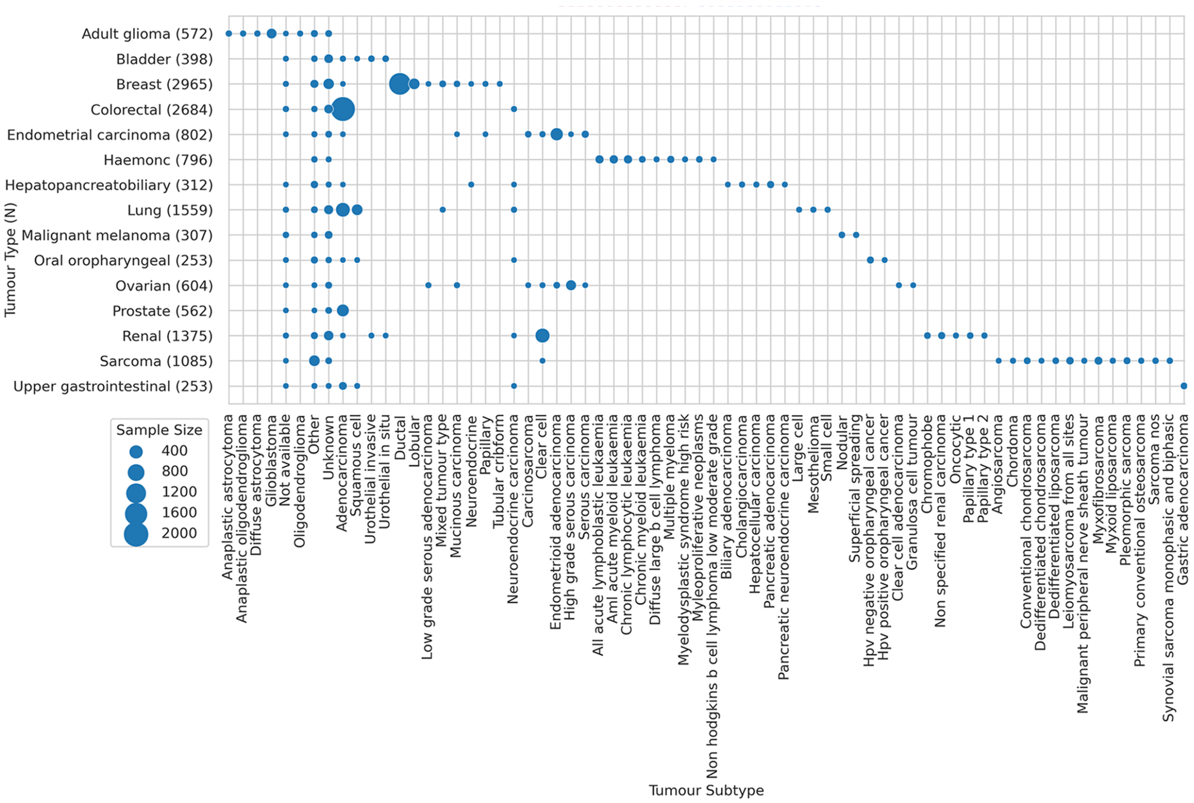


Supplementary Figure 1: Number of tumours in the Genomics England dataset per tumour type (Y-axis) and subtype (X-axis).

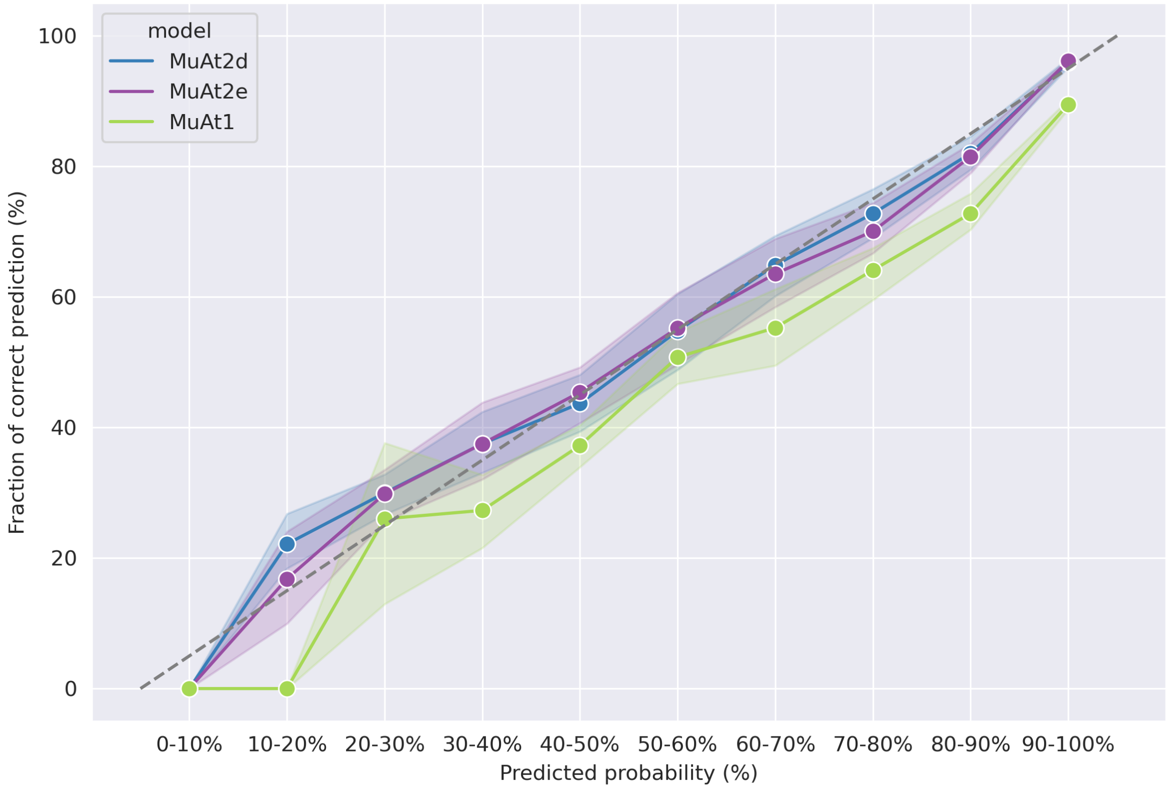


Supplementary Figure 2: Comparison of models calibration between MuAt1-pretrained on PCAWG and MuAt2-finetuned on GEL

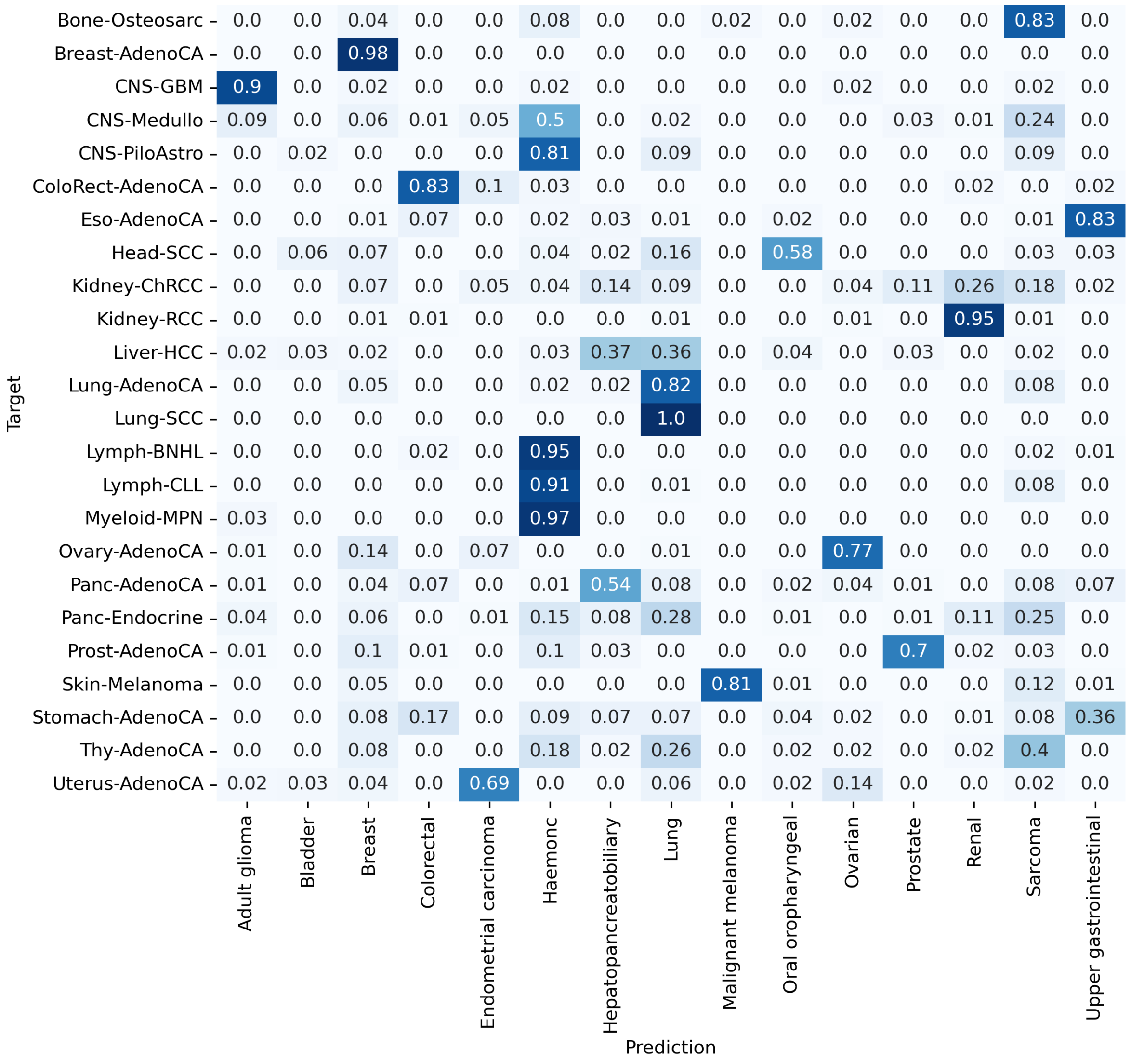


Supplementary Figure 3: MuAt2 external validation on PCAWG. Validation performed on an unseen PCAWG pretrained set to ensures unbiased predictive performance.

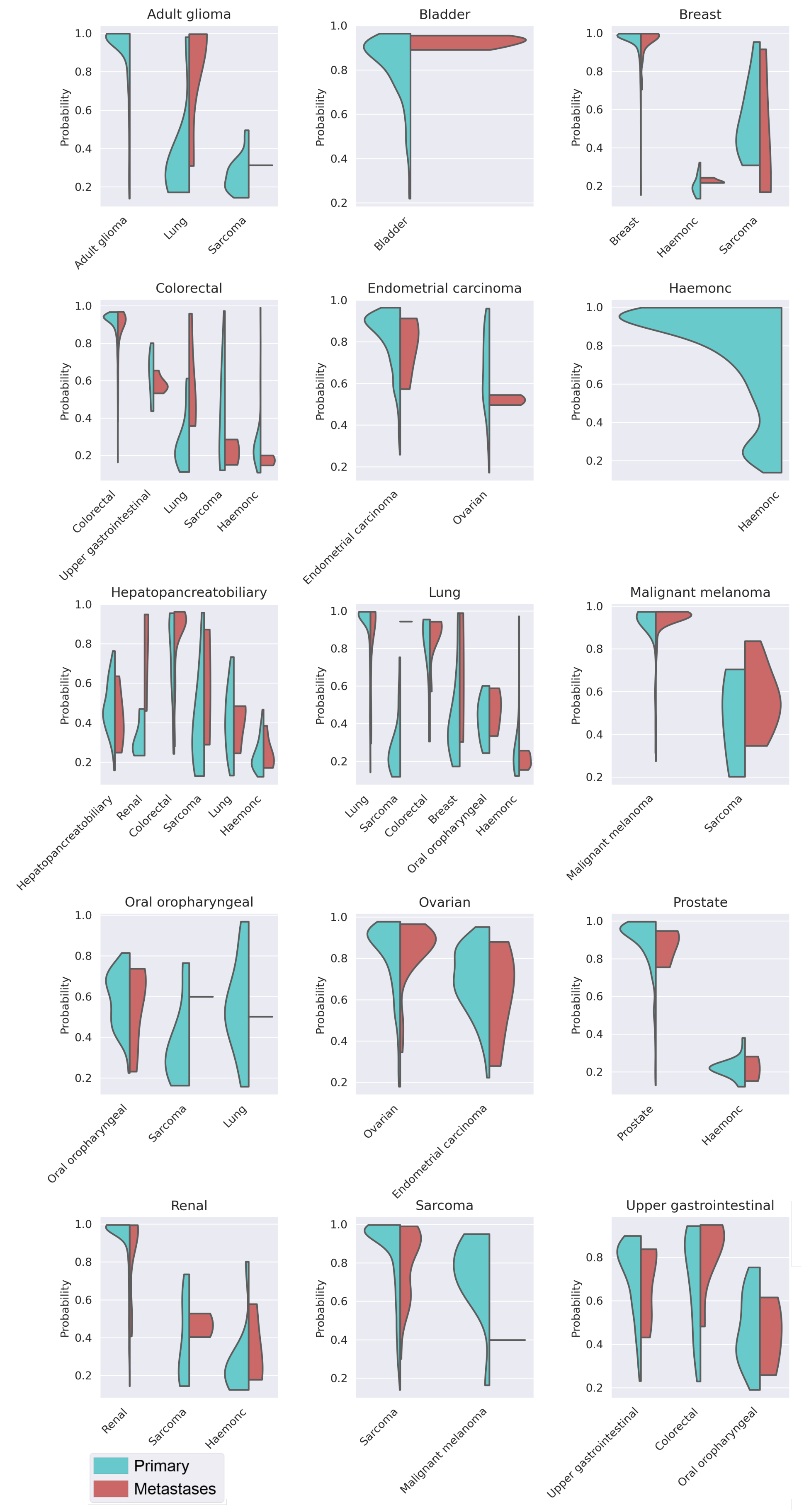


Supplementary Figure 4: top-1 probability distribution from MuAt2 prediction (x-axis).

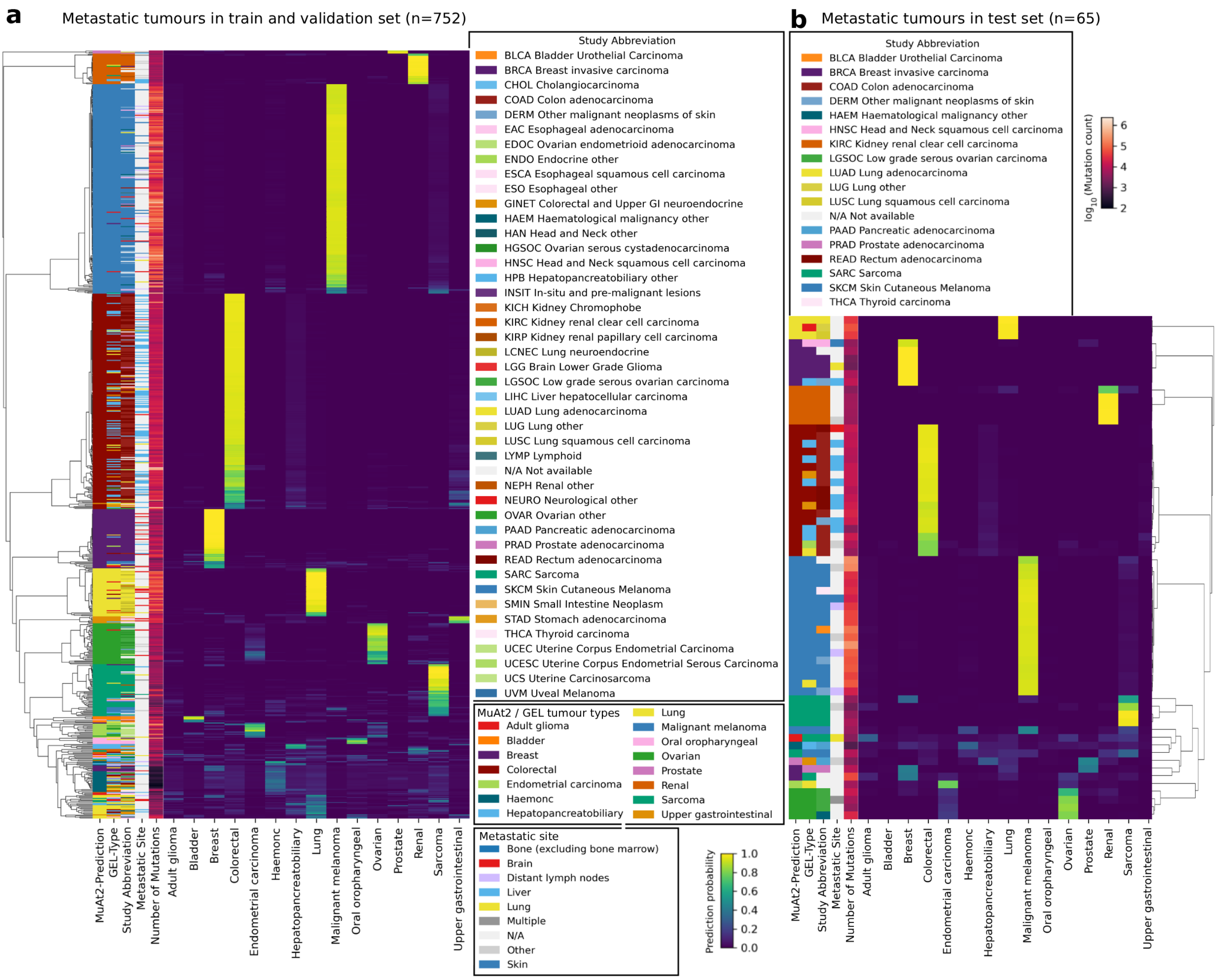


Supplementary Figure 5: Clustermap of MuAt2 prediction on metastatic tumours in **a**) training, validation set and **b**) test set.

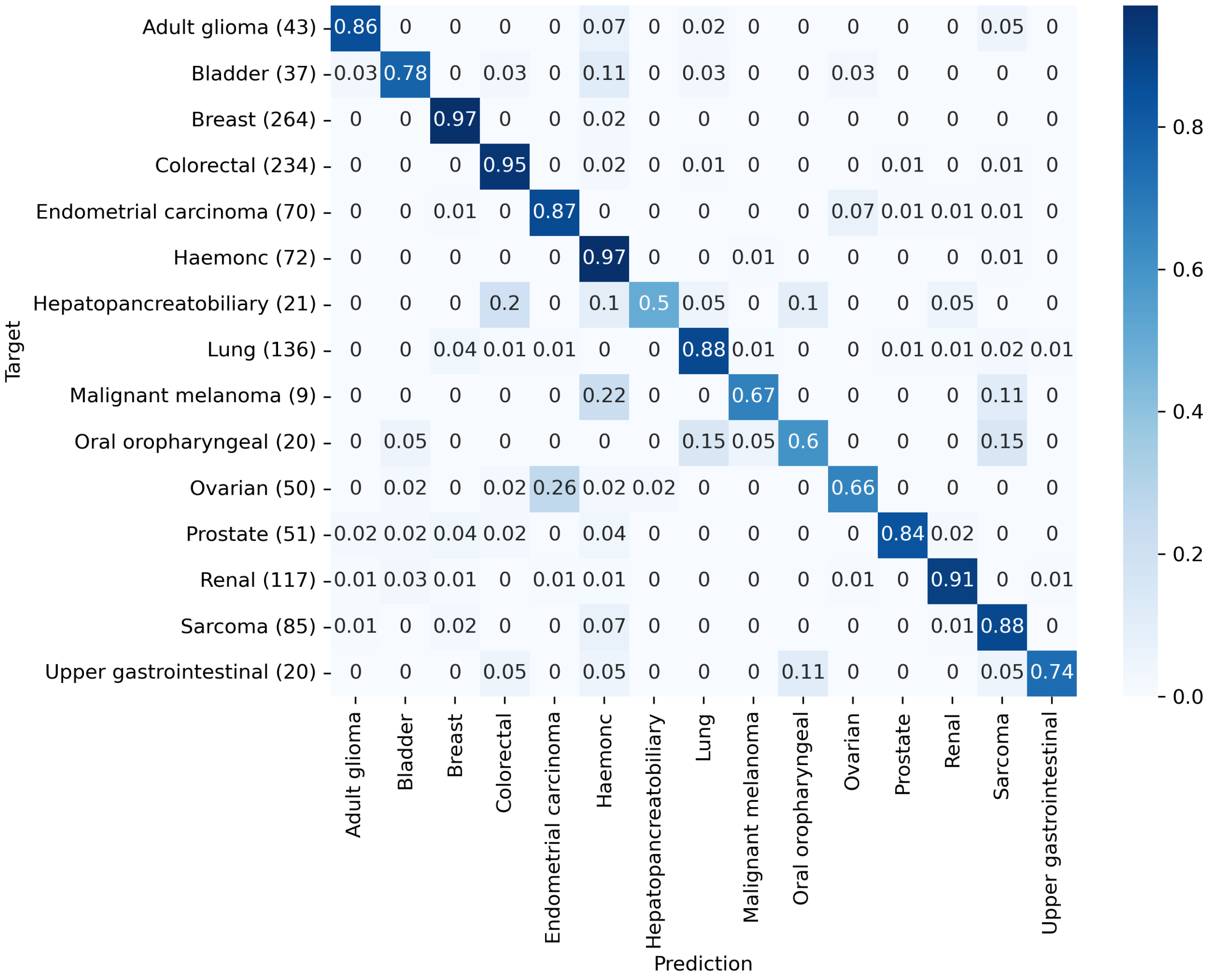


Supplementary Figure 6: Confusion matrix of MuAt2 prediction on tumours with no metastatic tags.

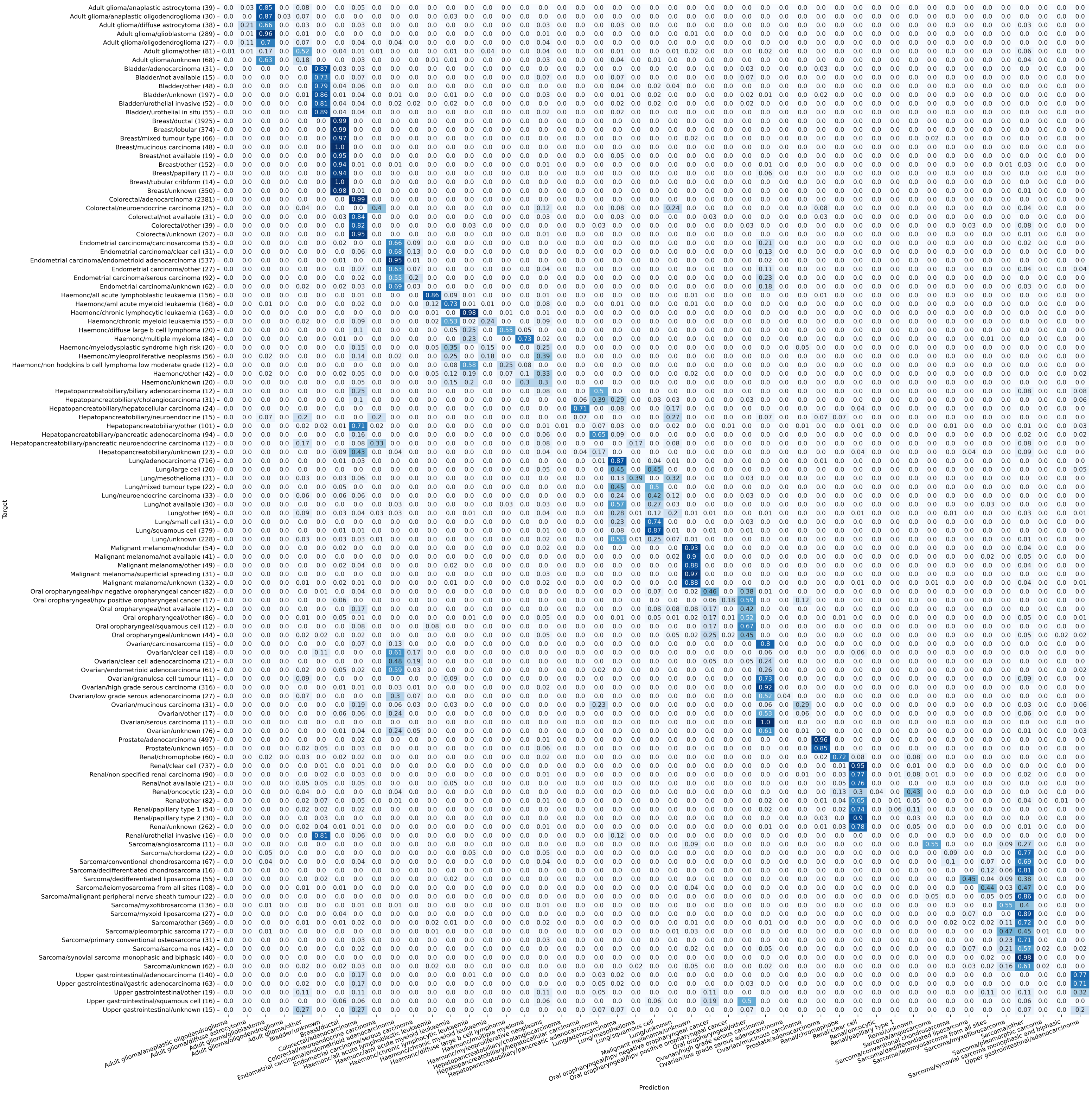


Supplementary Figure 7: Confusion matrix of MuAt2 combined type-subtypes in all GEL set.

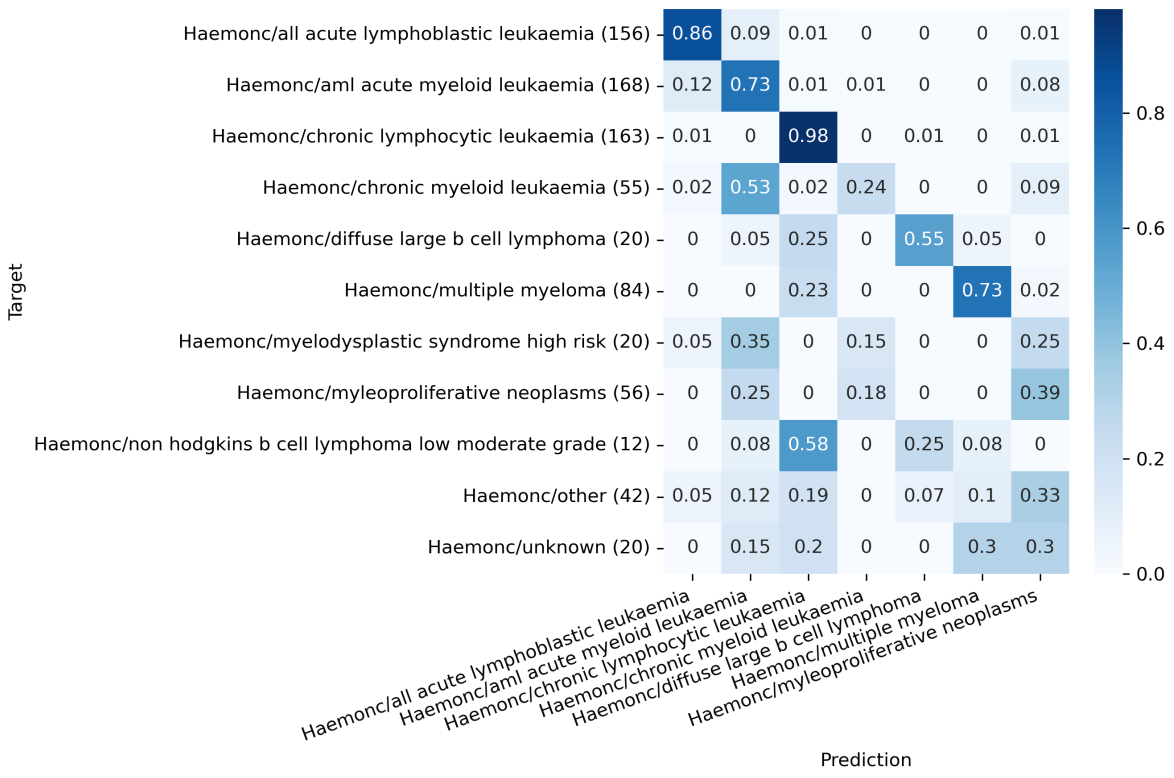


Supplementary Figure 8: Confusion matrix of haematological subtypes in all GEL data

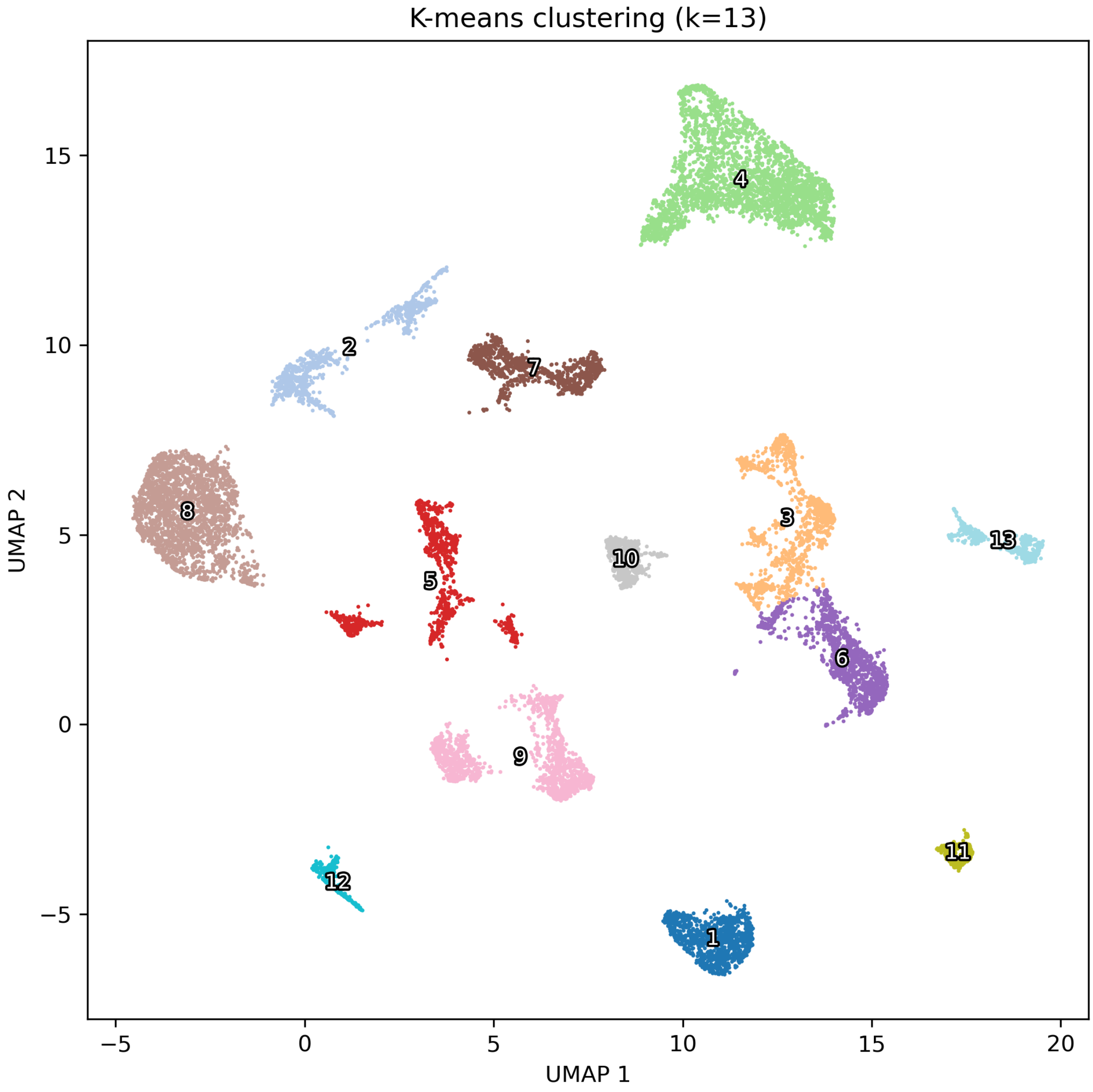


Supplementary Figure 9: Primary k-means clustering of MuAt2 latent features. UMAP projection of MuAt2 principal components coloured by the 13 primary k-means clusters selected using the silhouette score. Each cluster is labelled with its ID (1–13).

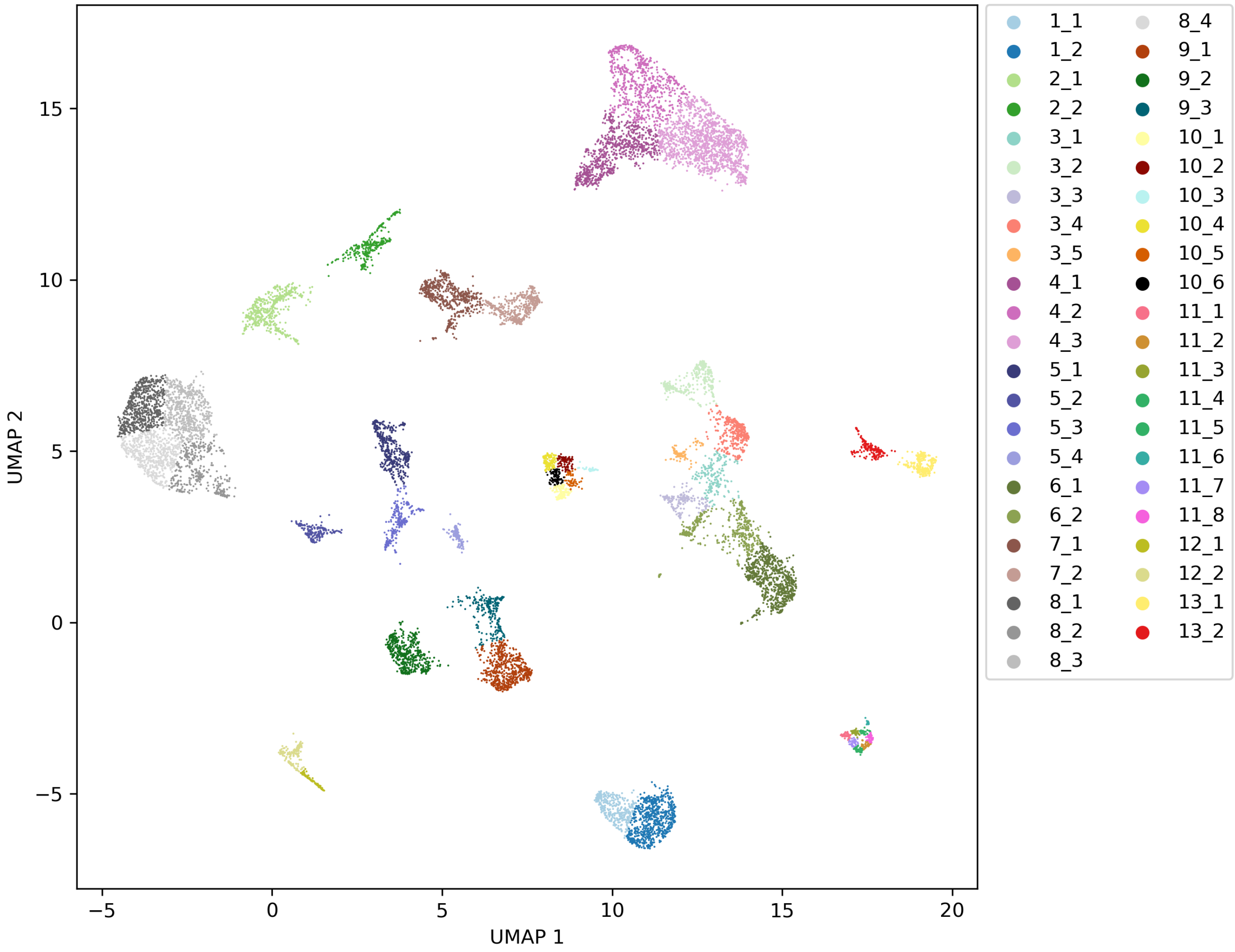


Supplementary Figure 10: First and second-level k-means clustering of MuAt2 latent features. UMAP projection of MuAt2 principal components coloured by subclusters obtained by reapplying k-means with the silhouette criterion within each of the 13 primary clusters. Each colour corresponds to one of the resulting 45 secondary clusters; the legend shows the primary cluster ID followed by the subcluster ID (e.g. "1_1"–"1_n").

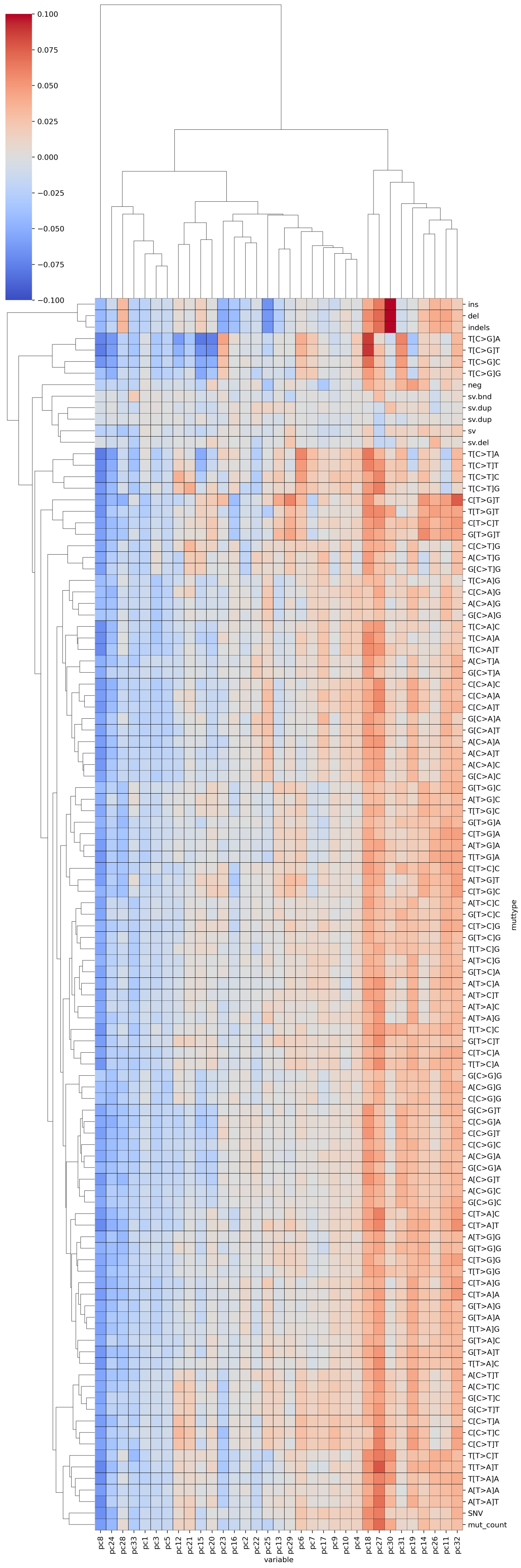




Supplementary Figure 11: Associations between MuAt2 principal components and mutational burden and mutation-type counts.

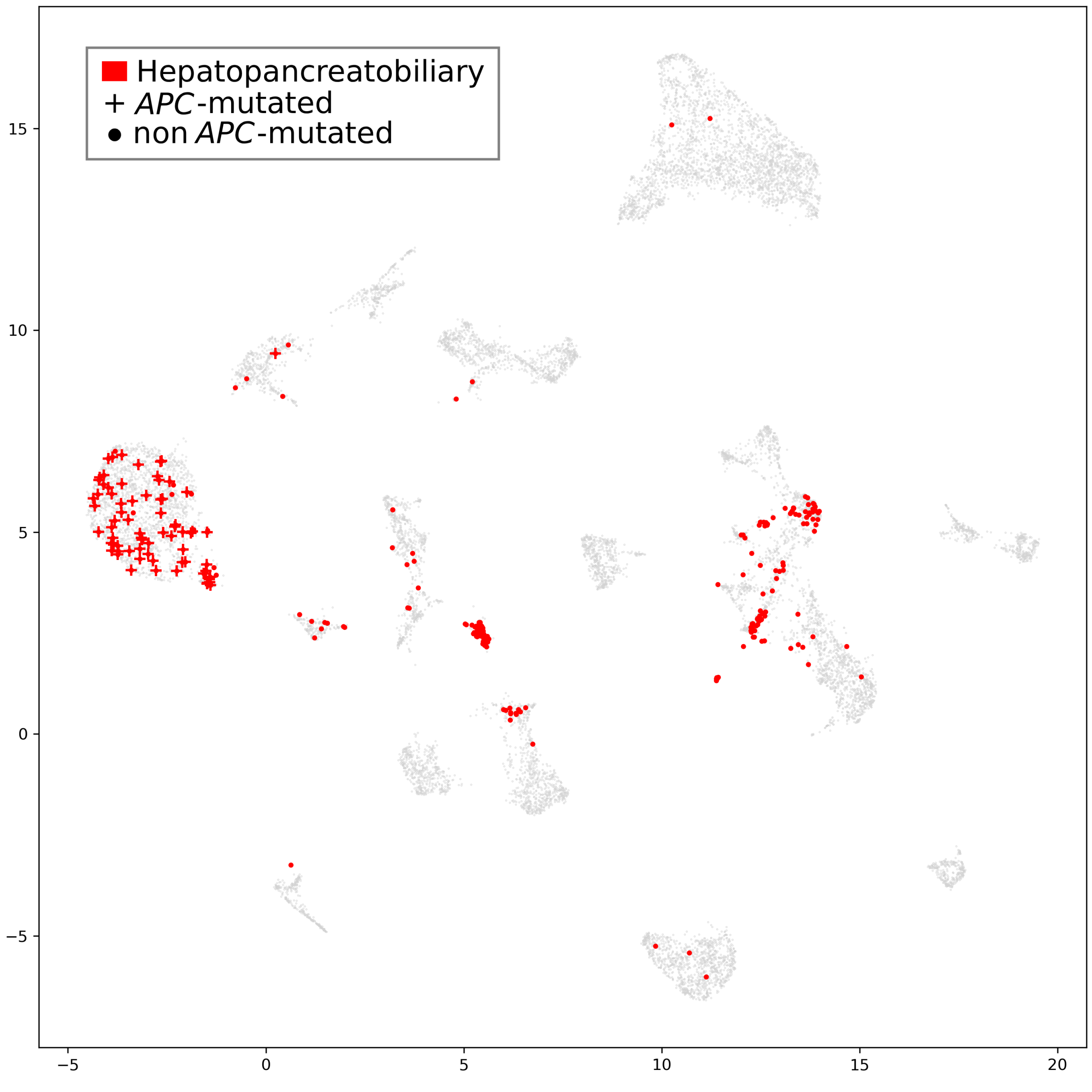


Supplementary Figure 12: Distribution of $APC$-mutated hepatopancreatobiliary tumours in the MuAt2 latent space.

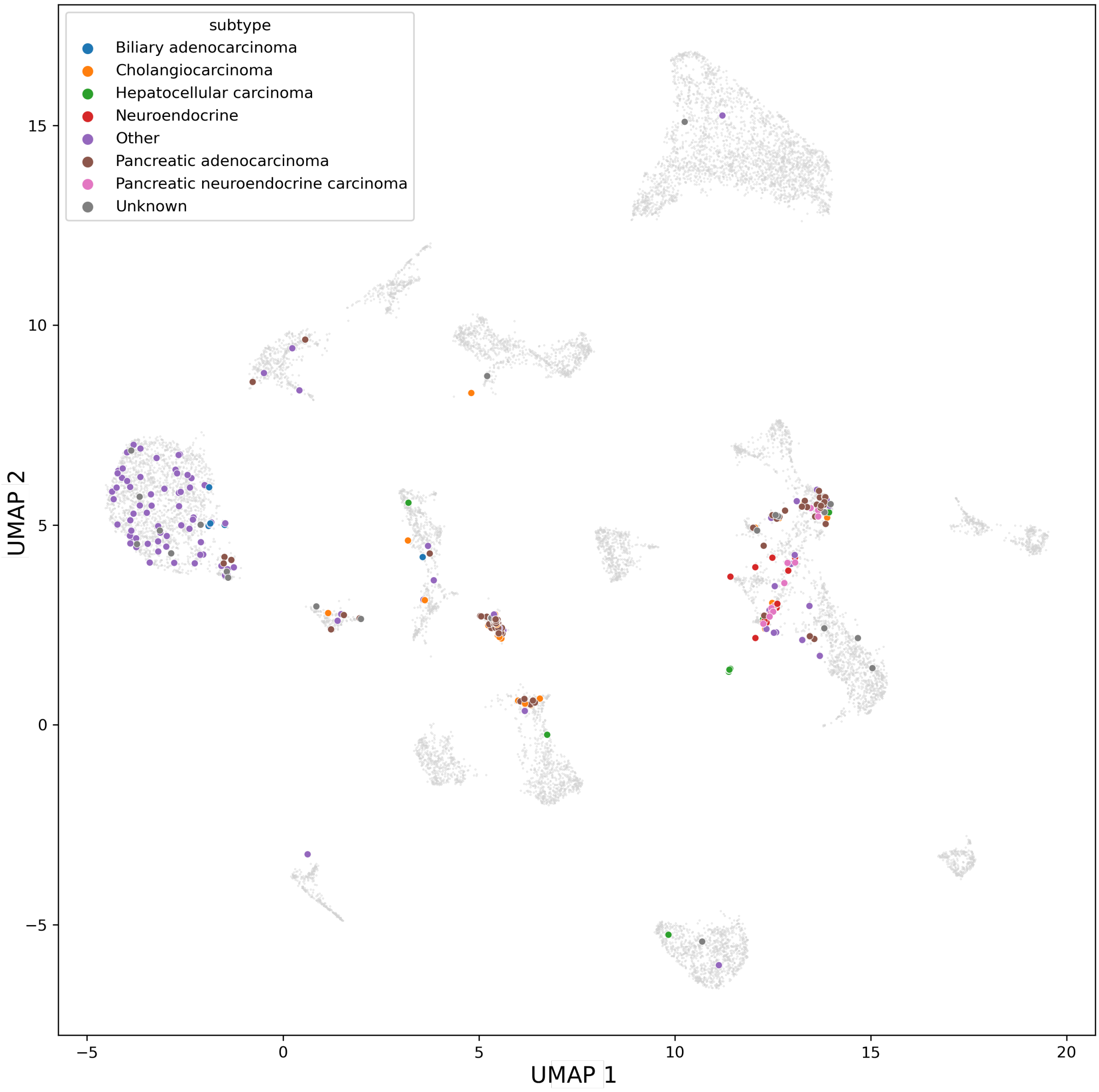


Supplementary Figure 13: Colorectal subtypes in the MuAt2 latent space.

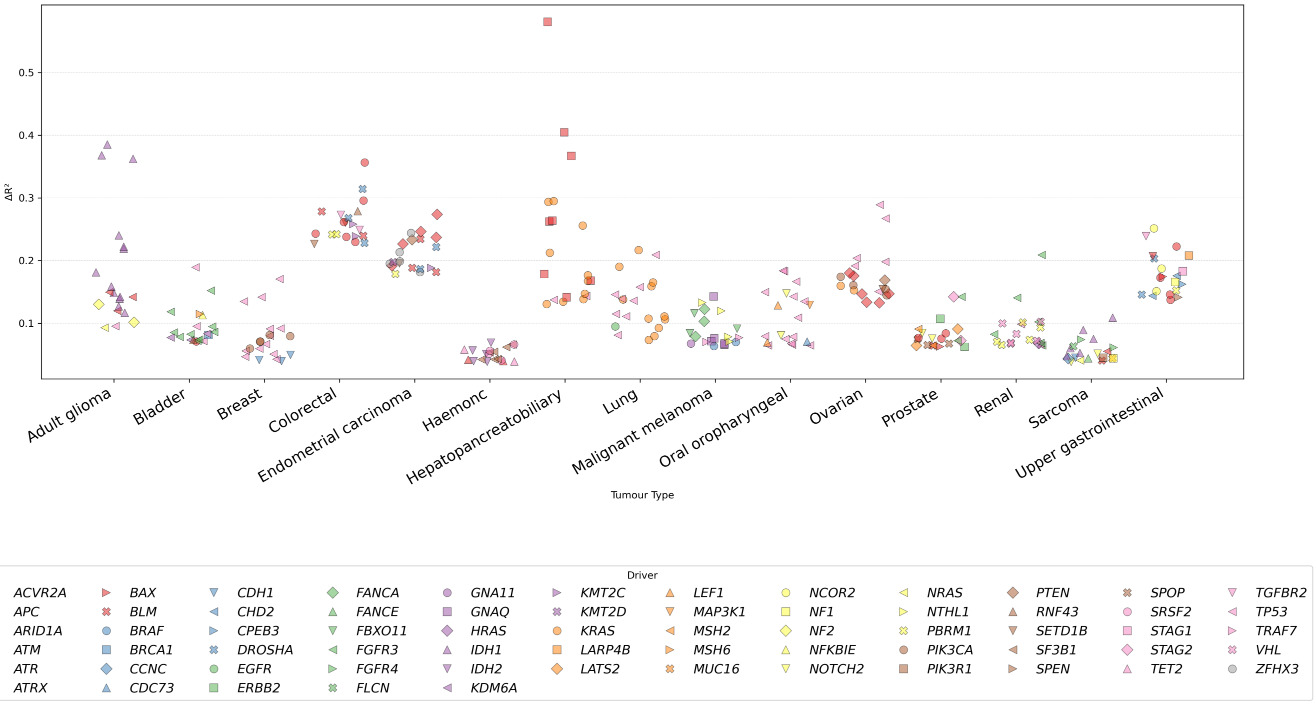


Supplementary Figure 14: Change in explained variance ($\Delta R^2$) of MuAt2 principal components by cancer driver events when including high TMB, MSI and *POLE*-ultramutated samples.

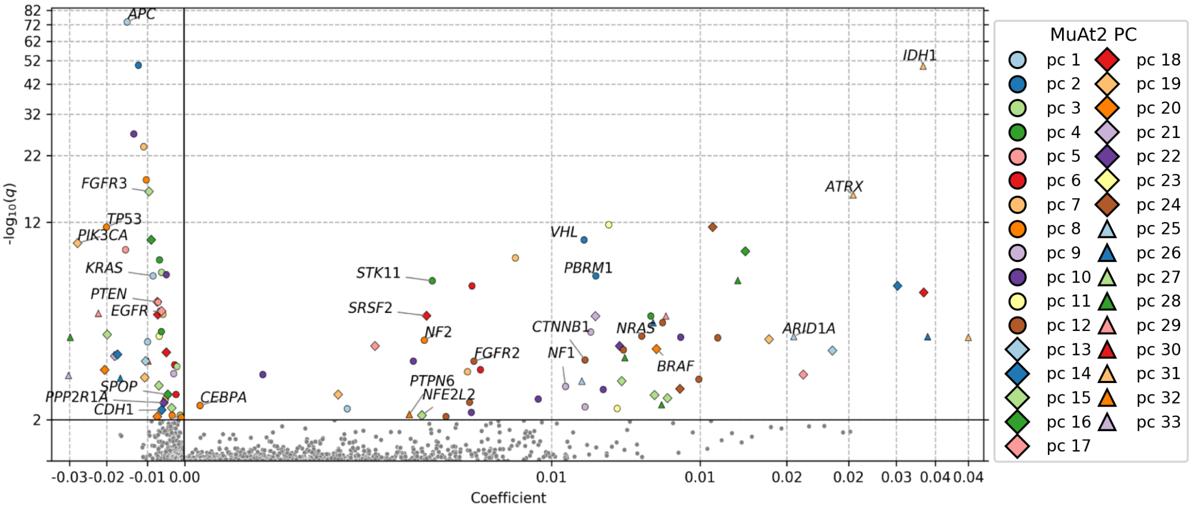


Supplementary Figure 15: Volcano plot of MuAt features associated with drivers in all tumours (pan-cancer). High TMB, MSI, and POLE samples were excluded from the analysis.

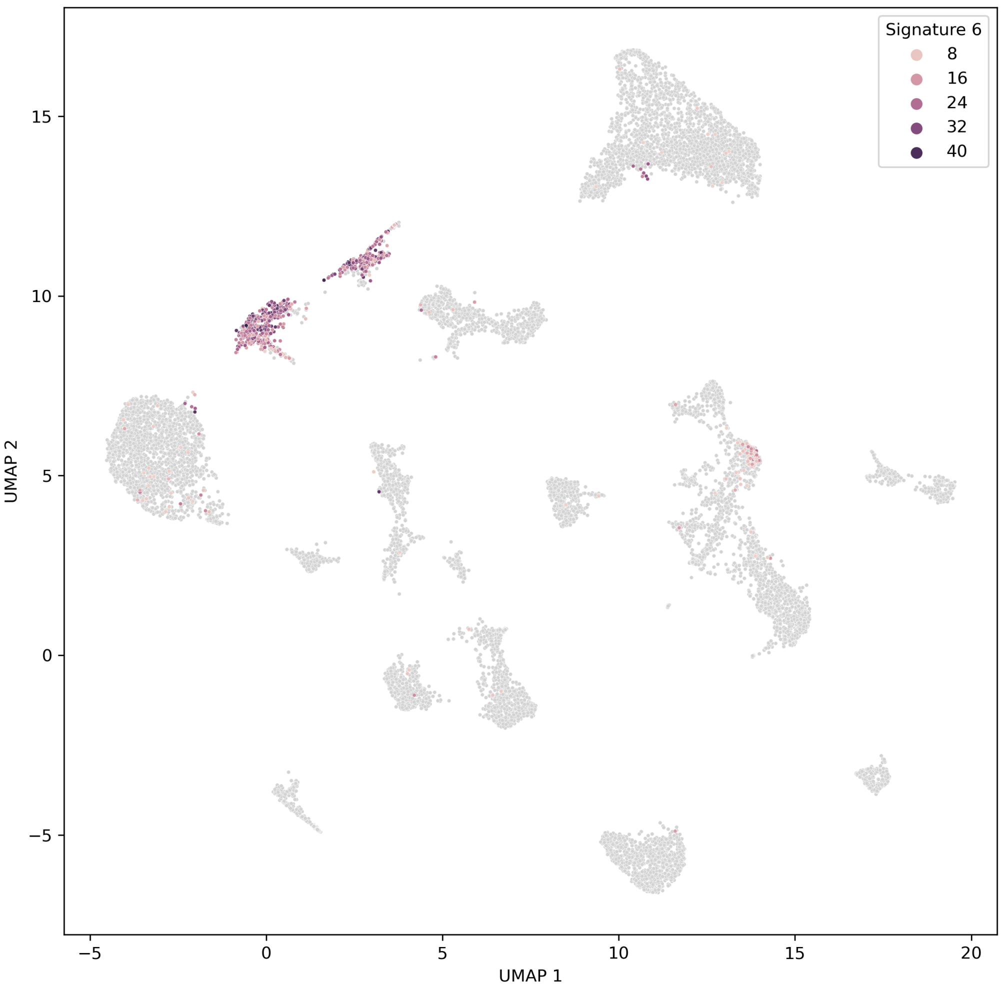


Supplementary Figure 16: Distribution of SBS6 mutational signature (MMR deficiency) exposure (%) across the MuAt2 latent space.

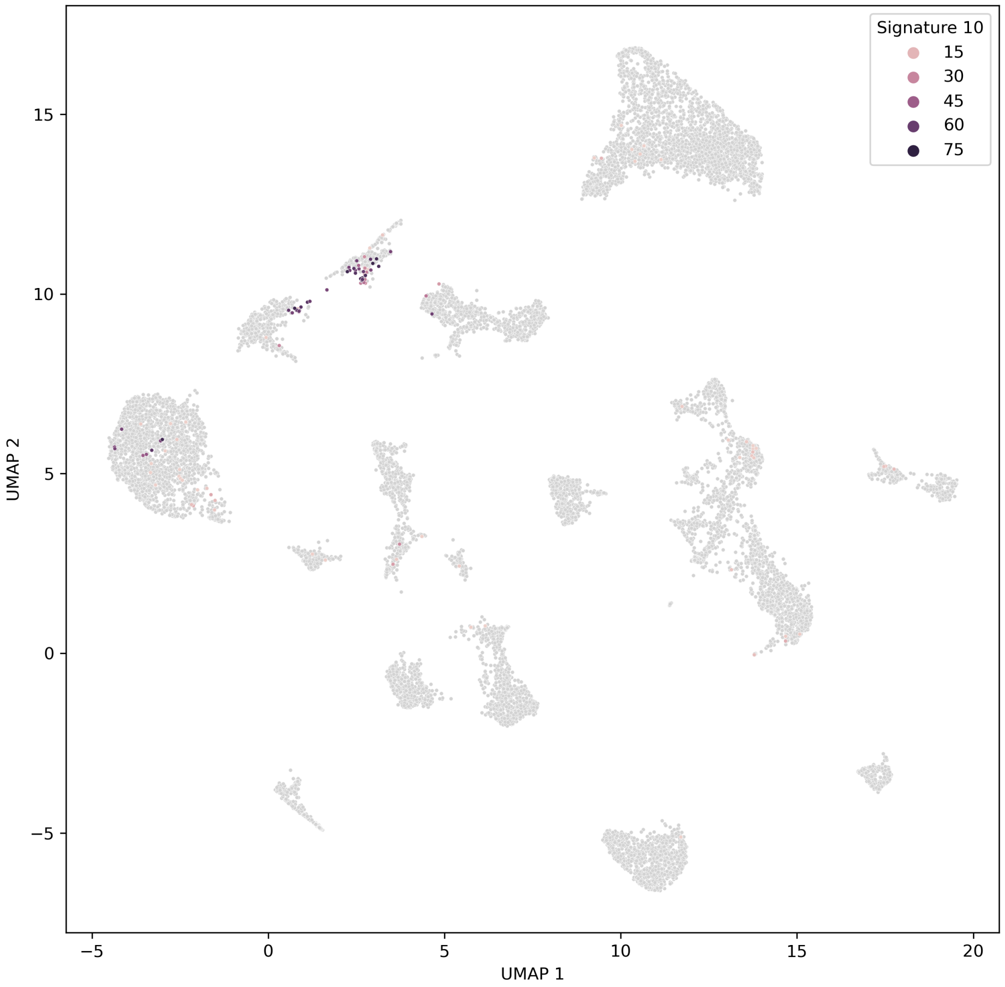


Supplementary Figure 17: Distribution of SBS10 mutational signature (*POLE*) exposure (%) across the MuAt2 latent space.

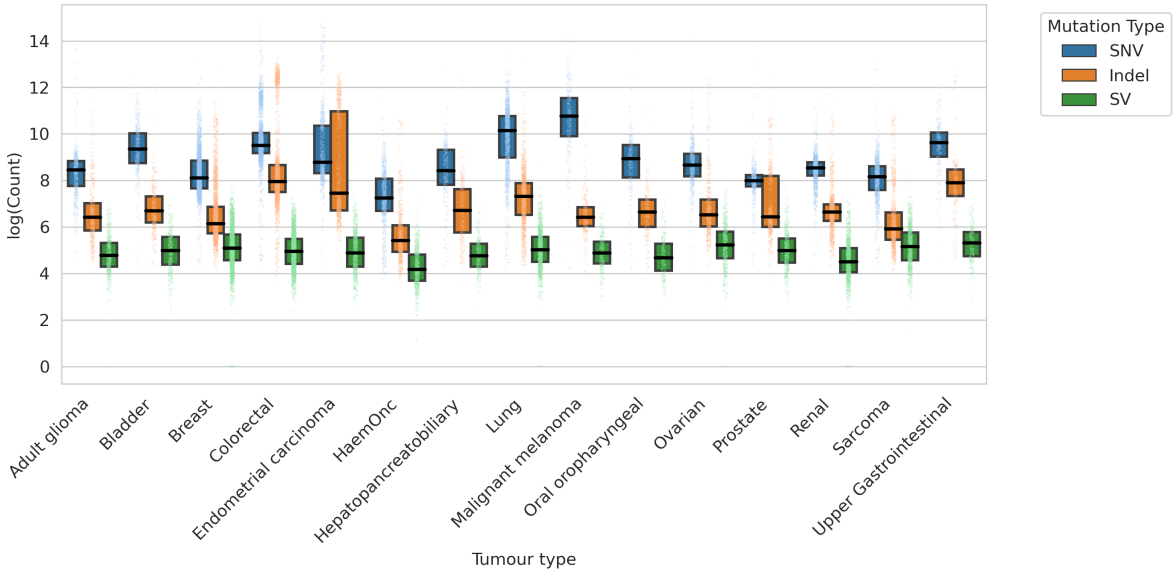


Supplementary Figure 18: Number of mutations in a log scale across tumour types.

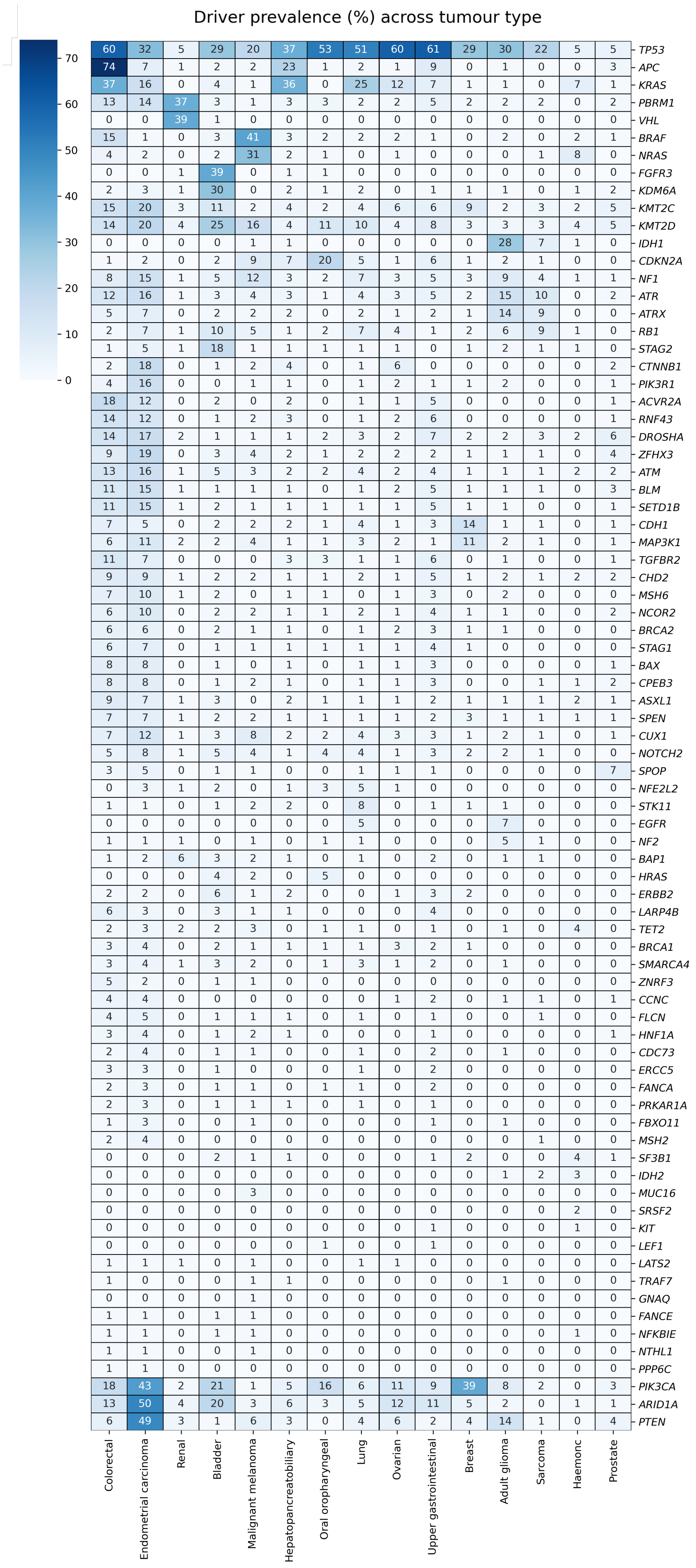




Supplementary Figure 19: Prevalence of known driver events across tumour types.

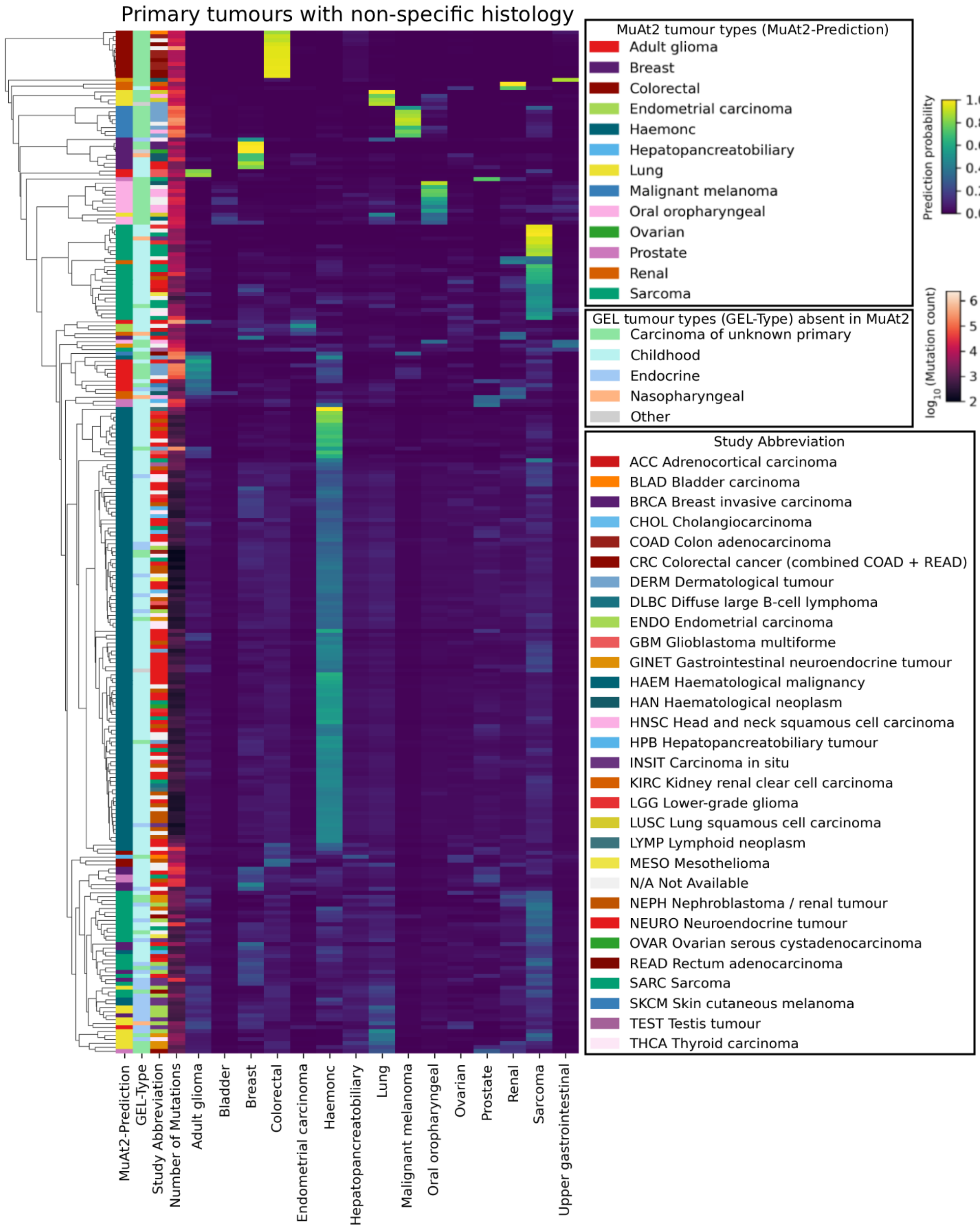


Supplementary Figure 20: MuAt2 prediciton on held-out primary tumours with non-specific histology.

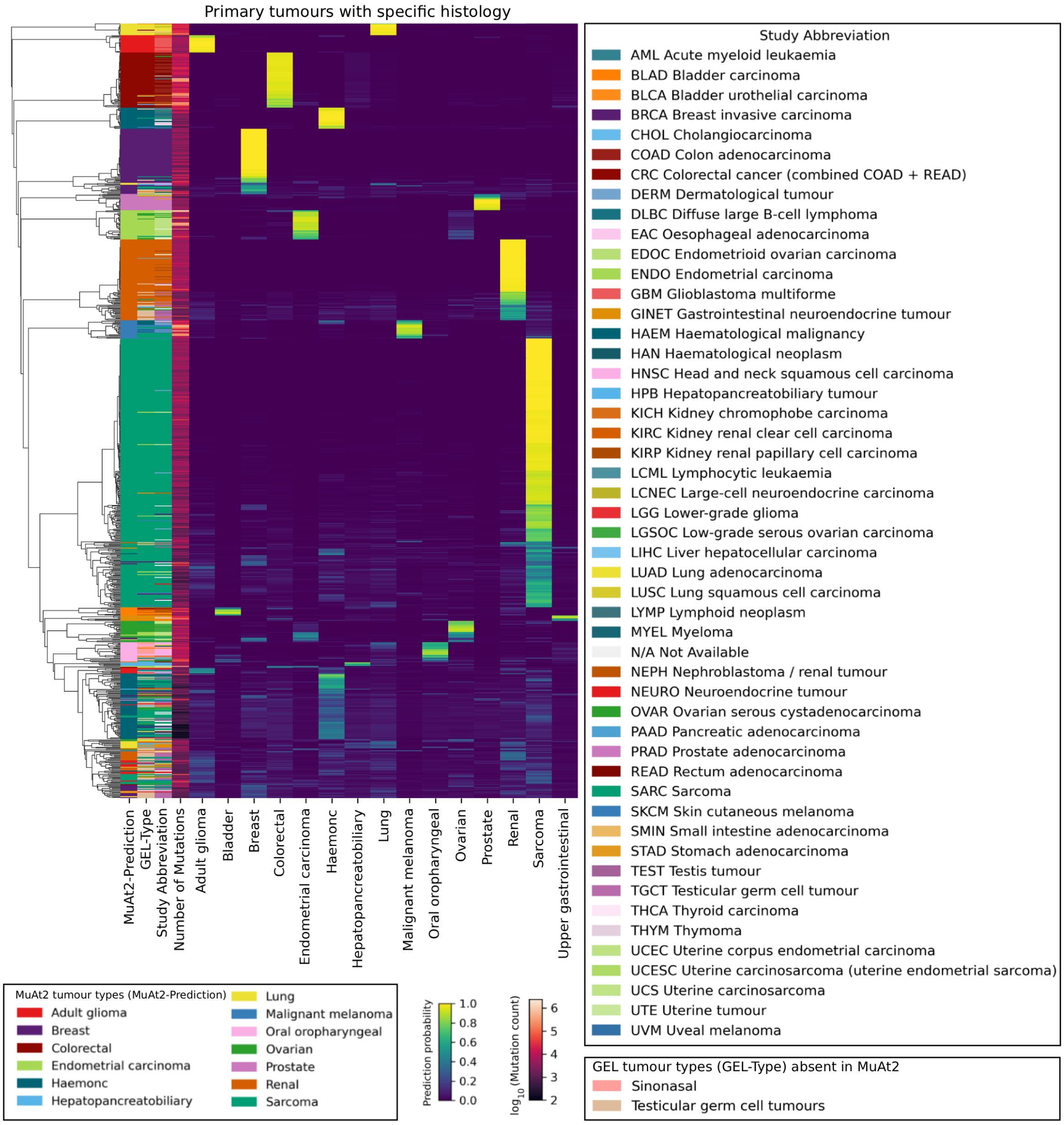


Supplementary Figure 21: MuAt2 prediciton on held-out primary tumours with specific histology.

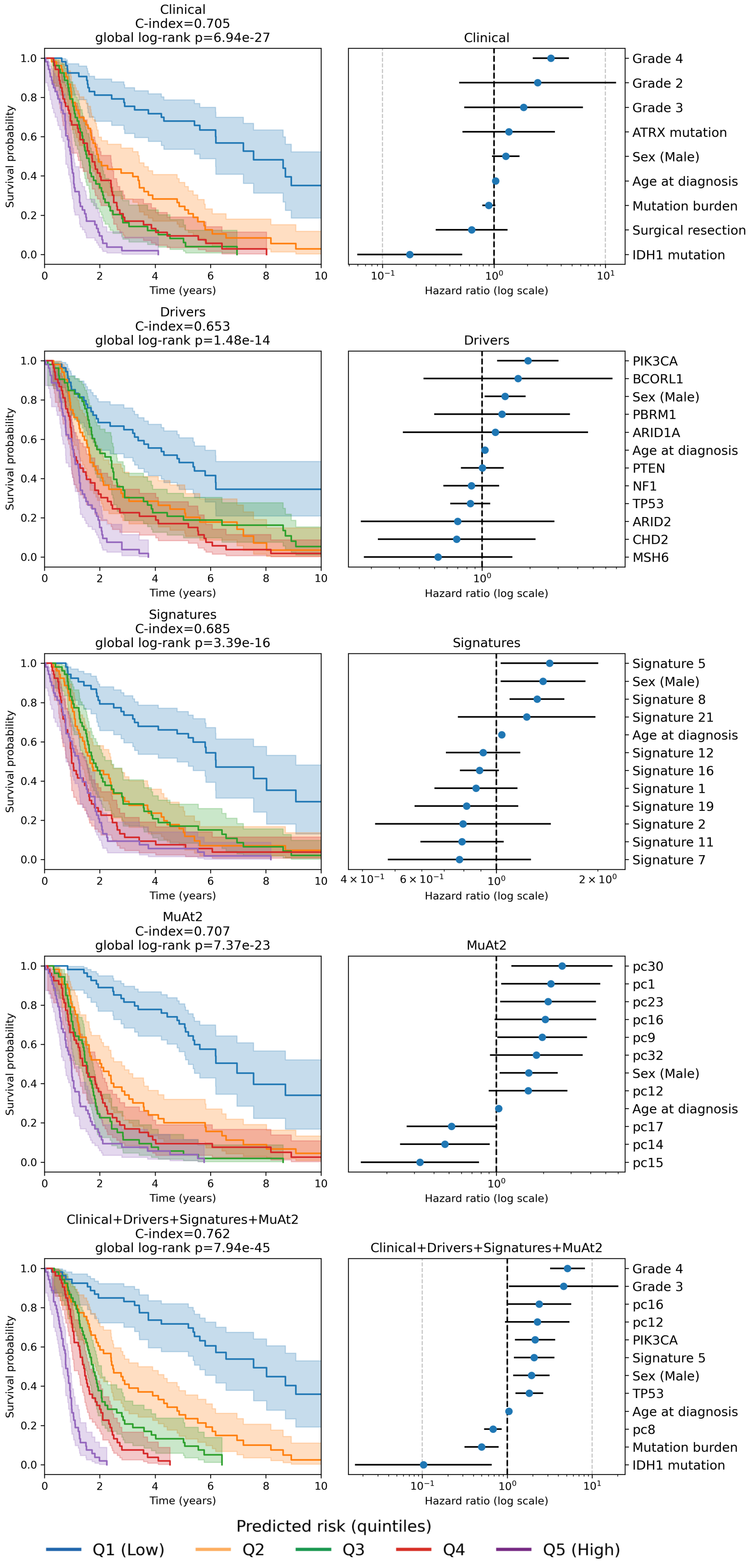


Supplementary Figure 22: Prognostic stratification in glioblastoma (n = 266). Risk-stratified survival and corresponding multivariable hazard ratios across clinical, driver, signature and MuAt2-based Cox models.

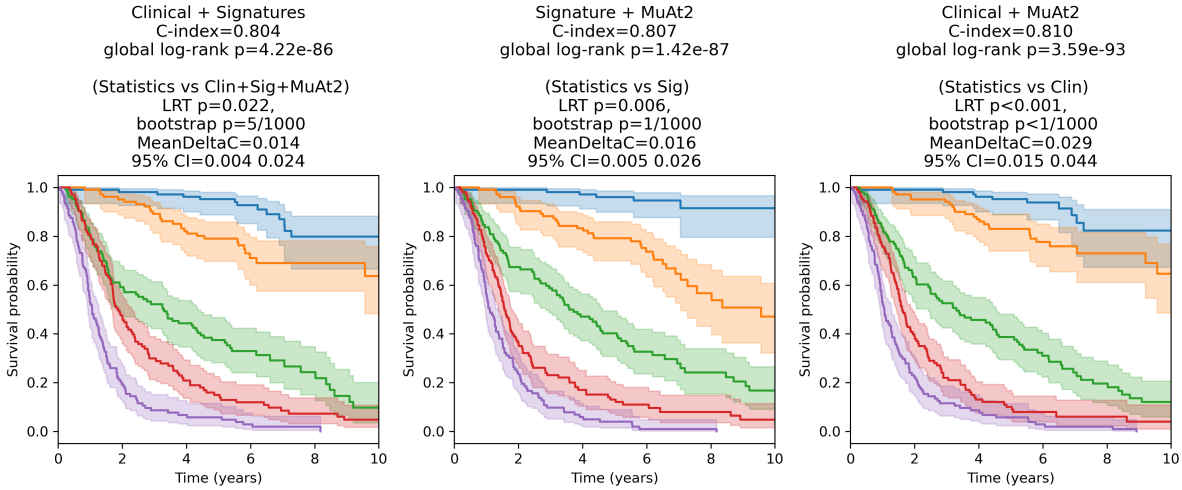


Supplementary Figure 23: Kaplan–Meier survival stratification in adult glioma(n=518) using Cox models based on combinations of clinical variables, mutational signatures, MuAt2 features. Patients were grouped into equal-sized predicted risk quintiles.

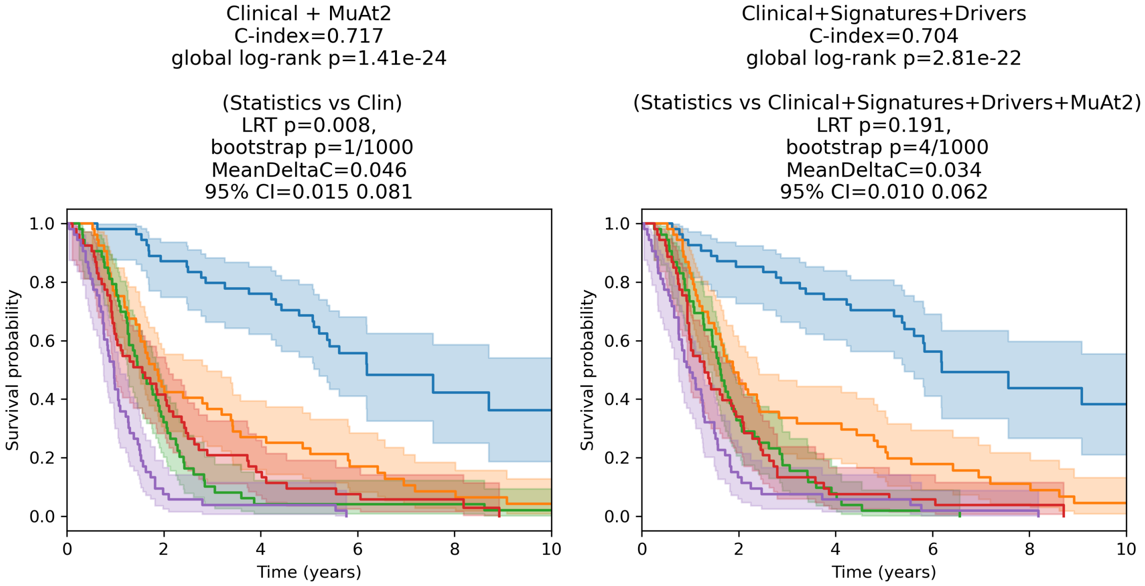


Supplementary Figure 24: Kaplan–Meier survival stratification in adult glioma(n=266) using Cox models based on combinations of clinical variables, mutational signatures, MuAt2 features. Patients were grouped into equal-sized predicted risk quintiles.
