## Supplementary tables for "Pan-cancer tumour classification and risk stratification from whole-genome somatic variants via dual-task representation learning": supplementary_tables.pdf

[3]iCAN Digital Precision Cancer Medicine Flagship, Finland

Supplementary Table 1: Predictive performance of MuAt2 and baseline models

Supplementary Table 2: Significant enrichments (FDR $< 10\%$) of driver events within MuAt2 primary $k$-means clusters (k=13).

Supplementary Table 3: Significant enrichments (FDR $< 10\%$) of driver events within MuAt2 secondary $k$-means clusters (k=45).

Supplementary Table 4: Change in explained variance ($\Delta R^2$) of MuAt2 principal components by cancer driver events when excluding high TMB, MSI and *POLE*-ultramutated samples.

Supplementary Table 5: Associations between MuAt2 principal components and mutation-type burden.

Supplementary Table 6: Change in explained variance ($\Delta R^2$) of MuAt2 principal components by cancer driver events when including high TMB, MSI and *POLE*-ultramutated samples.

Supplementary Table 7: Association analysis of MuAt2 principal components for individual driver events beyond tumour-type and subtype in low TMB, no POLE and no MSI tumours for each tumour types.

Supplementary Table 8: Genomic England dataset information

Supplementary Table 9: MuAt2 hyperparameters

Supplementary Table 10: Prevalence of known driver events across tumour types.

Supplementary Table 11: Association analysis between driver and mutation-type burden

Supplementary Table 12: Multivariable Cox survival analysis results across clinical, mutational signature, MuAt2 and combined models in glioma.

Supplementary Table 13: Multivariable Cox survival analysis results across clinical, mutational signature, MuAt2 and combined models in glioblastoma.
